# Tuberculosis disease severity assessment using clinical variables and radiology enabled by artificial intelligence

**DOI:** 10.1101/2024.08.19.24311411

**Authors:** Marwan Ghanem, Ratnam Srivastava, Yasha Ektefaie, Drew Hoppes, Gabriel Rosenfeld, Ziv Yaniv, Alina Grinev, Ava Y. Xu, Eunsol Yang, Gustavo E. Velásquez, Linda Harrison, Alex Rosenthal, Radojka M. Savic, Karen R. Jacobson, Maha R. Farhat

## Abstract

Radiology can define tuberculosis (TB) severity and may guide duration of treatment, however the optimal radiological metric to use and which clinical variables to combine it with in the real-world is unclear. We systematically associated baseline chest X-rays (CXR) metrics with TB treatment outcome using real-world data from diverse TB clinical settings. We used logistic regression to associate 10 radiological metrics including percent of lung involved in disease (PLI), cavitation, and Timika score, alone or with other clinical characteristics, stratifying by drug resistance and HIV (n = 2,809). We fine-tuned convolutional neural nets (CNN) to automate PLI measurement from the CXR DICOM images (n = 5,261). PLI is the only CXR finding associated with unfavorable outcome across drug resistance and HIV subgroups [rifampicin-susceptible disease without HIV, adjusted odds ratio 1·11 (1·01, 1·22), P-value 0·025]. The most informed model of baseline characteristics tested predicts outcome with a validation mean area under the curve (AUC) of 0·769. PLI alone predicts unfavorable outcomes equally or better than Timika or cavitary information (AUC PLI 0·656 vs. Timika 0·655 and cavitation best 0·591). PLI>25% provides a better separation of favorable and unfavorable outcomes compared to PLI>50% currently used in some clinical trials. The best performing ensemble of CNNs has an AUC 0·850 for PLI>25% and mean absolute error of 11·7% for the PLI value. PLI is better than cavitation, is accurately predicted with CNNs, and is optimally combined with age, sex, and smear grade for predicting unfavorable treatment outcome in pulmonary TB in real-world settings.

**Significance Statement:** A systematic evaluation of specific CXR findings in combination with clinical variables and their association with unfavorable outcomes in real-world settings is currently lacking. Stratification by severity of pulmonary TB can support personalized treatment, including the identification of patient groups that can be cured reliably with a shortened treatment regimen. Shorter regimens can minimize drug side effects, improve adherence and reduce costs of care. With the wider use of digital CXR and the increased adoption of AI for computer assisted diagnosis, radiology has the potential to be leveraged for multiple uses in the treatment and monitoring of TB disease, including contributing to a more individualized approach to TB treatment.

## Introduction

Pulmonary tuberculosis (TB) has a wide spectrum of clinical presentation ranging from incidentally-found asymptomatic disease to severe lung destruction with cachexia and multisystem organ failure(1). The extent of pulmonary disease and its secondary effects on other organ systems is thought to influence short term prognosis, treatment response and long-term sequelae of TB(2, 3). A standardized and generalizable measure of baseline severity for TB disease can support the optimization of treatment regimens, guide care resources for treatment monitoring, and prognostication(4, 5). Such a measure can inform clinical trial design and stratified enrollment for TB across the drug resistance spectrum. Although several measures are in current use to stratify patients based on severity profiles in clinical trials (5–8), these have varied significantly from study to study. In addition, the basis for the choice of these measures has been chosen based on either expert review only or analysis of small patient samples from a single site without external validation, or have utilized high-cost limited access tools such as PET-CT that are unlikely to be deployed in clinical practice(9–11). Specifically, associations of chest X-ray (CXR) findings with unfavorable outcomes or severity have commonly focused on the presence of lung cavitation(4–8, 12, 13). The Timika or Ralph score sums the percent of lung involved in disease (PLI) on CXR with 40 points added if any cavities are present(11). Other approaches to assessing radiological severity have included a count of the number of zones affected by disease (0-6)(12, 13) and a dichotomization of PLI at a threshold of 50%(7, 14, 15).

In addition to radiology, multiple clinical variables have been highlighted as associated with unfavorable treatment outcomes, including male sex(4), advanced age(4, 16), low BMI(4, 16), alcohol use(16), diabetes mellitus(17), malignancy(18), HIV co-infection(4), smear positivity or grade(4), and low adherence to treatment(4). It is not clear what the real-world value of CXR findings is for treatment response prediction or if radiological variables can improve treatment response prediction when combined with non-radiological variables. Here, we systematically study baseline CXR findings alone or in combination with other clinical variables and assess their association with TB treatment outcome. We used the TB-Portals database(19) that collects multimodal information from drug-susceptible and drug-resistant TB across geographically diverse real-world treatment settings. In conjunction, we aim to automate the measurement of the most predictive CXR findings using machine learning to facilitate access to severity assessment in high TB prevalence settings.

## Results

### Patient inclusion, baseline characteristics and treatment outcomes

At the time of access, TB-Portals included data on 11,282 care episodes (11,067 patients) from 13 countries between 2008-2023. The most well represented countries were Ukraine (n = 3,176), Georgia (n = 2,953), Moldova (n = 1,280) (**Table S1, Fig. S1**). 2,809 patient care episodes fit our inclusion criteria (**Fig. 1**). We stratified patients into three groups based on rifampicin susceptibility and HIV: (a) without HIV + rifampicin-susceptible TB: training-validation n = 566 (Rif-S1), test n = 285 (Rif-S2), (b) without HIV + rifampicin-resistant TB: training-validation n = 1,056 (Rif-R1), test n = 530 (Rif-R2), and (c) with HIV + any TB: n = 372. Compared with Rif-S1, the Rif-R1 and HIV subgroups had a higher frequency of prior TB, anemia, smoking, alcohol use, and other comorbidities (**Table 1**). People with HIV had a higher frequency of extrapulmonary disease, low BMI, smoking, alcohol use, and drug use than people without HIV. Pulmonary nodules (Rif-S1 81%) and cavities (Rif-S1 37%) were the most common CXR findings across all groups. The Rif-R1 group had the highest frequency of cavities (45%), and people with HIV the lowest (28%). Cavities were most commonly small (<3 cm) (all groups, Rif-S1 24%). Median PLI ranged from 18% to 26%, and Timika from 26 to 45 across the three groups (**Table 1**).

**Figure 1.**
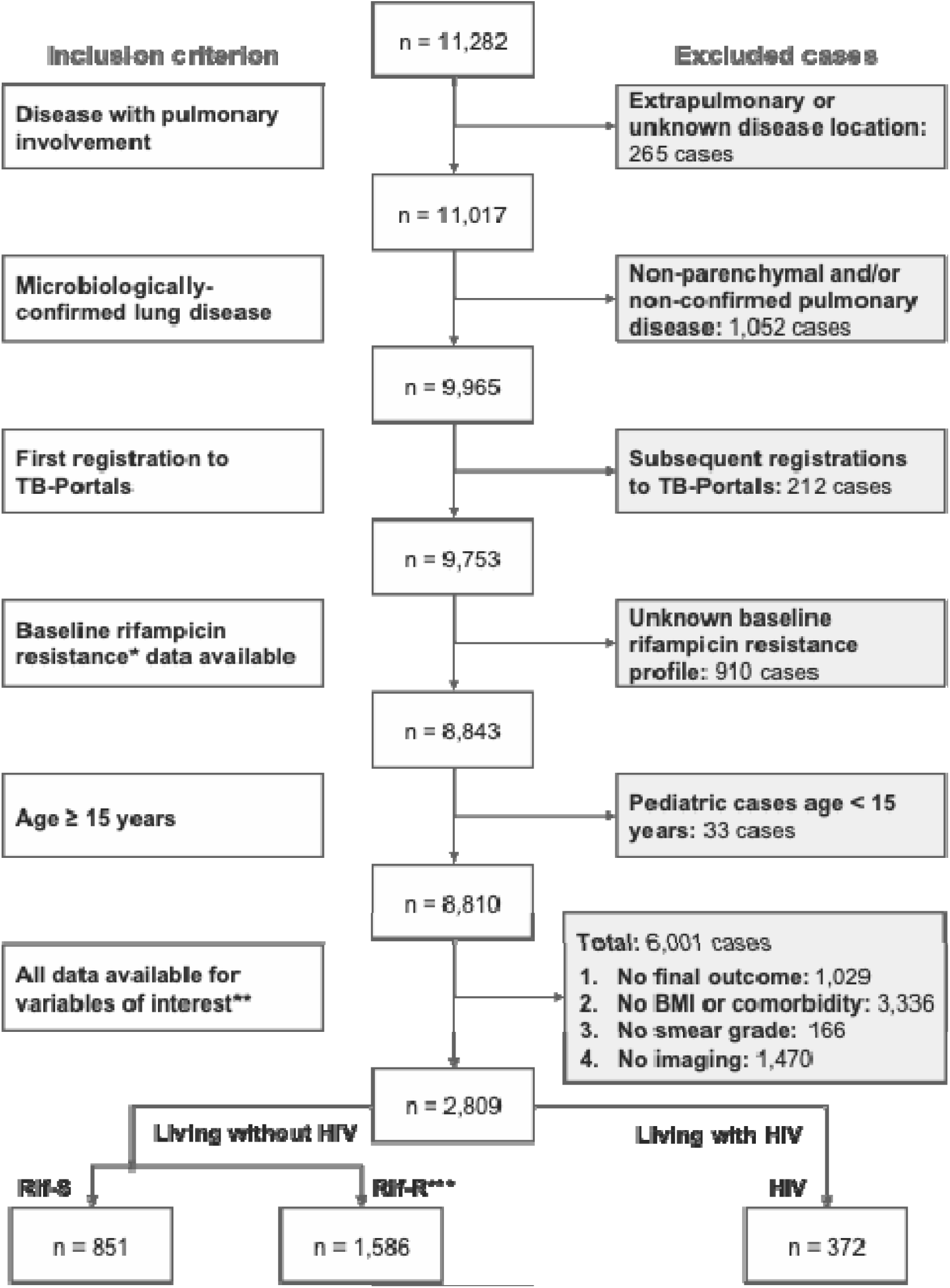
Inclusion criteria. We queried TB Portals for patients with lung involvement, microbiologically-confirmed disease, first registration to TB Portals, Age at onset ≥15 years, and all data available for features of interest and outcomes. *Baseline rifampicin resistance: between 90 days pre-treatment initiation and 30 days post-treatment initiation, inclusive. Rifampicin susceptibility testing is done through one or more of the following tests: BACTEC MGIT 960, Lowenstein-Jensen, Line-Probe Assay, Truenat, or GeneXpert. **Removal of missing data for variables of interest was done in a stepwise manner: final outcome (“cured”, “completed”, “failure”, “died” and “palliative care” as non-missing final outcome), BMI and comorbidities, smear-grade and culture data, chest X-ray data. ***Rifampicin resistance includes isolates that tested as resistant or intermediate on drug susceptibility testing (phenotypic or genotypic).

**Table 1.** Baseline patient characteristics and final outcome. Frequencies are shown as No. (%) and continuous variables are shown as median (IQR). *Other comorbidity includes hepatic or renal disease, diabetes mellitus, immunosuppression, pneumoconiosis or other diseases. **Effective TB drugs include 4+ regimen RIPE or equivalent for ≥60 days or until death, or 4+ second-line regimen for ≥150 days or until death. ***unfavorable outcomes include treatment failure, death or palliative care. Rif-S1 = people living without HIV + rifampicin-susceptible TB, Rif-R1 = people living without HIV + rifampicin-resistant TB, PLI = percent of lung involved in disease, lymphadenopathy = mediastinal lymphadenopathy

| Features | Living without HIV (n = 2,437) |  |  | Living with HIV (n = 372) |
| --- | --- | --- | --- | --- |
|  | Rif-S1 (n = 566) | Rif-R1 (n = 1,056) | Test dataset (n = 815) |  |
| Baseline characteristics |  |  |  |  |
| Female | 140 (25%) | 291 (28%) | 210 (26%) | 100 (27%) |
| Age at onset (years) | 44 (34, 54) | 42 (32, 52) | 44 (35, 56) | 41 (36, 46) |
| BMI ≤ 18 kg/m<sup>2</sup> | 169 (30%) | 329 (31%) | 304 (37%) | 147 (40%) |
| Prior TB | 88 (16%) | 403 (38%) | 260 (32%) | 137 (37%) |
| Country |  |  |  |  |
| Most common | Moldova (48%) | Ukraine (46%) | Ukraine (85%) | Ukraine (68%) |
| Second most common | Ukraine (29%) | Belarus (17%) | Belarus (8%) | Moldova (15%) |
| Other | Other (23%) | Other (37%) | Other (7%) | Other (17%) |
| Extrapulmonary | 16 (3%) | 55 (5%) | 12 (1%) | 32 (9%) |
| Comorbidity |  |  |  |  |
| Anemia | 20 (4%) | 104 (10%) | 141 (17%) | 75 (20%) |
| Other | 151 (27%) | 464 (44%) | 397 (49%) | 151 (41%) |
| Smoking | 148 (26%) | 544 (52%) | 521 (64%) | 256 (69%) |
| Alcohol use | 86 (15%) | 253 (24%) | 294 (36%) | 141 (38%) |
| Drug use | 6 (1%) | 31 (3%) | 36 (4%) | 86 (23%) |
| Smear grade |  |  |  |  |
| Scanty (vs. 0) | 89 (16%) | 135 (13%) | 134 (16%) | 63 (17%) |
| 1+ (vs. 0) | 109 (19%) | 231 (22%) | 181 (22%) | 69 (19%) |
| ≥2+ (vs. 0) | 143 (26%) | 281 (27%) | 259 (32%) | 97 (26%) |
| Rifampin resistance | 0 (0%) | 1,056 (100%) | 530 (65%) | 298 (80%) |
| Antiretrovirals | 0 (0%) | 0 (0%) | 0 (0%) | 65 (17%) |
| Effective TB regimen** | 481 (85%) | 728 (69%) | 689 (85%) | 289 (78%) |

| Baseline radiological features |  |  |  |  |
| --- | --- | --- | --- | --- |
| **Timika** | 45 (15, 80) | 45 (12, 69) | 40 (10, 68) | 26 (8, 60) |
| **Percent of lung involved in disease (PLI)** | 26 (11, 54) | 19 (9, 38) | 19 (10, 38) | 18 (8, 40) |
| **Cavities** |  |  |  |  |
| Presence | 212 (37%) | 471 (45%) | 352 (43%) | 106 (28%) |
| Small | 138 (24%) | 375 (34%) | 301 (37%) | 91 (24%) |
| Medium | 89 (16%) | 164 (16%) | 73 (9%) | 24 (6%) |
| Large | 52 (9%) | 93 (9%) | 42 (5%) | 8 (2%) |
| Multiple | 90 (16%) | 171 (16%) | 120 (15%) | 40 (11%) |
| **Lymphadenopathy** | 153 (27%) | 181 (17%) | 275 (34%) | 108 (29%) |
| **Nodules (presence)** | 461 (81%) | 920 (87%) | 749 (92%) | 334 (90%) |
| **Pleural effusion** | 105 (19%) | 121 (11%) | 74 (9%) | 71 (19%) |
| **Treatment outcomes** |  |  |  |  |
| **Unfavorable outcome\*\*\*** | 85 (15%) | 205 (19%) | 211 (26%) | 149 (40%) |

### Non-radiological features associated with unfavorable outcomes

We built logistic regression models of treatment outcomes using 13 demographic, clinical, microbiological, and regimen variables for the Rif-S1 (n = 566), Rif-R1 (n = 1,056) and HIV subgroups (n = 372) (*complete* models, **Table 2, Table S2**). For people with HIV, in addition to the 13 variables we included rifampicin resistance and antiretroviral therapy. We identified high smear grade (≥ 2+) compared with smear-negative disease [Rif-S1 aOR 3.84 (1.94, 7.59), p-value <0.001] to be associated with unfavorable outcome in all three groups. Other features associated with unfavorable outcome were low BMI, older age at onset of disease, prior TB, smoking, alcohol use, anemia, low smear grade (scanty, or 1+ *vs.* smear negative disease), rifampicin resistance, and the lack of an effective TB regimen (**Table 2**).

**Table 2.** Baseline non-radiological features and their association with unfavorable outcomes. Unadjusted = univariate logit model, Adjusted = multivariate logit model adjusted for the following characteristics: female (male as referent), age at onset of disease (continuous), BMI ≤ 18 kg/m^2^ (>18 kg/m^2^ as referent), extrapulmonary involvement, prior TB disease, anemia, other comorbidities (includes hepatic or renal disease, diabetes mellitus, immunosuppression, pneumoconiosis or other diseases), smoking, alcohol use, smear grade (scanty, 1+ or ≥ 2+ with negative microscopy as a referent), and effective drug therapy, *i.e.* RIPE or equivalent ≥ 60 days or second-line 4+ antimicrobials ≥ 150 days without known resistance to any of the components. Rifampicin resistance and antiretroviral use are added to the adjustment for people living with HIV.

| Feature | Unadjusted OR<br>(95% CI) | P-value (OR) | Adjusted OR<br>(95% CI) | P-value<br>(aOR) |
| --- | --- | --- | --- | --- |
| **People living without HIV with rifampin-susceptible TB (Rif-S1, n = 566)** |  |  |  |  |
| Female | 0.45 (0.24, 0.86) | 0.016 | 0.51 (0.25, 1.04) | 0.064 |
| Age at onset (years) | 1.03 (1.01, 1.04) | 0.002 | 1.04 (1.02, 1.06) | <0.001 |
| BMI $\leq 18 \text{ kg/m}^2$ | 1.52 (0.94, 2.45) | 0.090 | 1.40 (0.82, 2.37) | 0.217 |
| Prior TB | 2.03 (1.16, 3.54) | 0.013 | 1.65 (0.88, 3.07) | 0.117 |
| Extrapulmonary | 1.32 (0.37, 4.72) | 0.673 | 1.92 (0.46, 7.99) | 0.372 |
| Smoking | 1.90 (1.17, 3.09) | 0.010 | 1.25 (0.70, 2.26) | 0.452 |
| Alcohol use | 1.93 (1.10, 3.40) | 0.022 | 1.47 (0.78, 2.79) | 0.233 |
| Anemia | 3.23 (1.25, 8.35) | 0.015 | 2.45 (0.78, 7.66) | 0.125 |
| Other comorbidity | 1.34 (0.81, 2.21) | 0.251 | 0.92 (0.51, 1.67) | 0.792 |
| **Smear grade (vs. negative)** |  |  |  |  |
| Scanty | 1.07 (0.57, 1.99) | 0.838 | 2.30 (1.03, 5.12) | 0.041 |
| 1+ | 1.70 (1.00, 2.90) | 0.050 | 3.37 (1.62, 7.01) | 0.001 |
| $\geq 2+$ | 2.14 (1.32, 3.48) | 0.002 | 3.84 (1.94, 7.59) | <0.001 |
| Effective TB regimen | 0.60 (0.34, 1.08) | 0.087 | 0.59 (0.31, 1.11) | 0.103 |
| **People living without HIV with rifampin-resistant TB (Rif-R1, n = 1,056)** |  |  |  |  |
| Female | 0.37 (0.24, 0.56) | <0.001 | 0.62 (0.39, 1.00) | 0.051 |
| Age at onset (years) | 1.04 (1.03, 1.05) | <0.001 | 1.04 (1.02, 1.05) | <0.001 |
| BMI $\leq 18 \text{ kg/m}^2$ | 1.67 (1.22, 2.29) | 0.001 | 1.56 (1.08, 2.24) | 0.017 |
| Prior TB | 2.10 (1.54, 2.86) | <0.001 | 1.59 (1.14, 2.24) | 0.007 |
| Extrapulmonary | 1.30 (0.69, 2.48) | 0.417 | 1.03 (0.50, 2.14) | 0.932 |
| Smoking | 2.20 (1.60, 3.03) | <0.001 | 1.15 (0.77, 1.70) | 0.497 |
| Alcohol use | 2.96 (2.14, 4.10) | <0.001 | 1.70 (1.15, 2.51) | 0.008 |
| Anemia | 4.76 (3.12, 7.26) | <0.001 | 2.85 (1.73, 4.69) | <0.001 |
| Other comorbidity | 1.67 (1.23, 2.26) | 0.001 | 1.33 (0.94, 1.88) | 0.113 |
| **Smear grade (vs. negative)** |  |  |  |  |
| Scanty | 0.94 (0.59, 1.49) | 0.778 | 1.50 (0.86, 2.61) | 0.153 |
| 1+ | 0.97 (0.67, 1.41) | 0.874 | 1.25 (0.79, 1.99) | 0.346 |
| $\geq 2+$ | 1.86 (1.34, 2.57) | <0.001 | 1.62 (1.05, 2.48) | 0.028 |
| Effective TB regimen | 0.47 (0.34, 0.64) | <0.001 | 0.44 (0.31, 0.63) | <0.001 |

| People living with HIV with any TB (HIV, n = 372) |  |  |  |  |
| --- | --- | --- | --- | --- |
| **Female** | 0.79 (0.49, 1.27) | 0.334 | 0.78 (0.45, 1.34) | 0.367 |
| **Age at onset (years)** | 1.01 (0.99, 1.04) | 0.266 | 1.02 (0.99, 1.05) | 0.170 |
| **BMI ≤ 18 kg/m<sup>2</sup>** | 2.34 (1.52, 3.59) | <0.001 | 2.47 (1.50, 4.08) | <0.001 |
| **Prior TB** | 1.34 (0.87, 2.06) | 0.179 | 1.03 (0.62, 1.70) | 0.920 |
| **Extrapulmonary** | 2.36 (1.13, 4.94) | 0.023 | 2.05 (0.85, 4.92) | 0.109 |
| **Smoking** | 1.08 (0.69, 1.69) | 0.738 | 0.56 (0.31, 0.98) | 0.043 |
| **Alcohol use** | 2.19 (1.43, 3.36) | <0.001 | 2.56 (1.51, 4.34) | 0.001 |
| **Anemia** | 2.26 (1.35, 3.78) | 0.002 | 2.11 (1.12, 3.96) | 0.020 |
| **Other comorbidity** | 1.07 (0.70, 1.64) | 0.743 | 0.81 (0.49, 1.33) | 0.397 |
| **Smear grade (vs. negative)** |  |  |  |  |
| Scanty | 0.77 (0.44, 1.35) | 0.362 | 1.06 (0.53, 2.14) | 0.869 |
| 1+ | 0.88 (0.52, 1.52) | 0.656 | 0.81 (0.41, 1.61) | 0.554 |
| ≥ 2+ | 2.38 (1.49, 3.81) | <0.001 | 2.17 (1.20, 3.92) | 0.011 |
| **Rifampin resistance** | 2.44 (1.37, 4.35) | 0.002 | 2.13 (1.08, 4.18) | 0.028 |
| **Antiretrovirals** | 0.92 (0.53, 1.60) | 0.773 | 0.58 (0.29, 1.14) | 0.116 |
| **Effective TB regimen** | 0.32 (0.19, 0.53) | <0.001 | 0.31 (0.17, 0.53) | <0.001 |

### Percent lung involved in disease (PLI) is associated with unfavorable TB treatment outcome

We studied ten radiological variables for association with unfavorable outcomes (**Table S2**). Supplementary Figure 2 shows examples of CXR images with low and high PLI, with and without cavitation. We added each variable one-by-one to the complete logistic regression models and used the Wald test for hypothesis testing of the coefficient (**Table 3**). PLI was significantly associated with unfavorable outcomes in all three groups [Rif-R1 group aOR 1.21 (1.13, 1.30) per 10% increase, p-value <0.001]. Timika was associated with unfavorable outcome in the Rif-R and HIV subgroups [Rif-R1 aOR 1.14 (1.08, 1.20) per ten-point increase, p-value <0.001]. Four cavitation variables were associated with unfavorable outcome in the Rif-R1 group (**Table 3**). The cavitation variable with the largest effect size was large cavities aOR 3.21(1.93, 5.33). Cavitation also improved model fit when added to a PLI-containing model for the Rif-R1 group (LRT p-value 0.016) (**Table S3**).

**Table 3.**
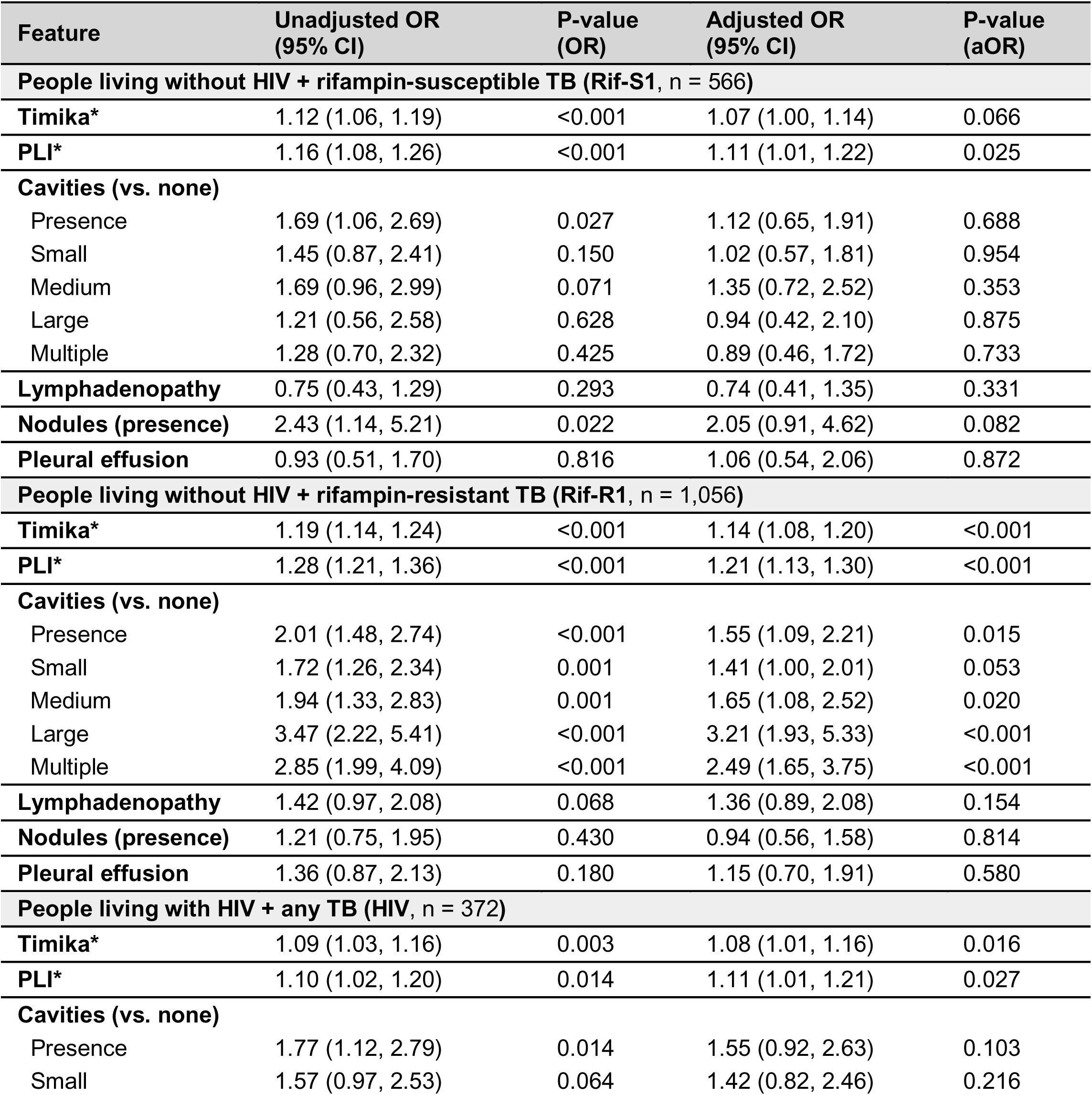
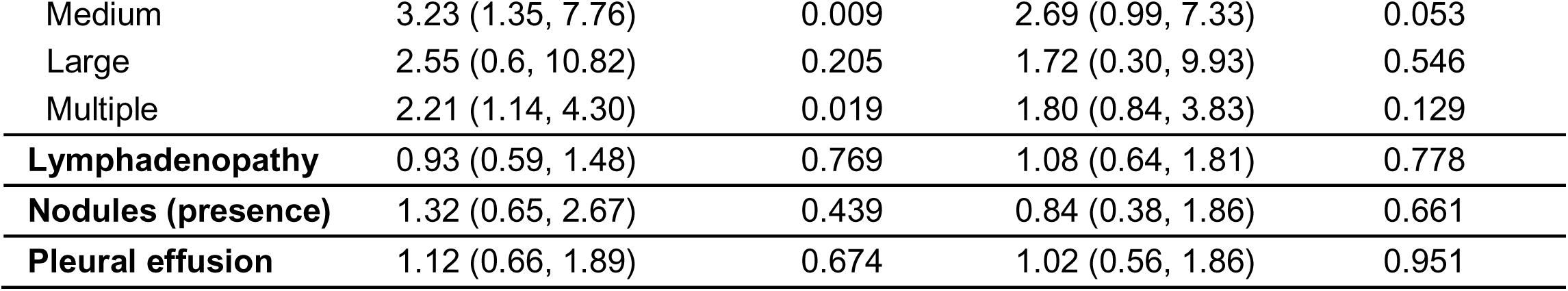
Baseline radiological features association with unfavorable outcomes. Unadjusted = univariate logit model, adjusted = multivariate logit model adjusted for the following characteristics: female (male as referent), age at onset of disease (continuous), BMI ≤ 18 kg/m^2^ (>18 kg/m^2^ as referent), extrapulmonary involvement, prior TB disease, anemia, other comorbidities (includes hepatic or renal disease, diabetes mellitus, immunosuppression, pneumoconiosis or other diseases), smoking, alcohol use, smear grade (scanty, 1+ or ≥ 2+ with negative microscopy as a referent), and effective drug therapy, *i.e.* RIPE or equivalent ≥ 60 days or second-line 4+ antimicrobials ≥ 150 days without known resistance to any of the components. Rifampicin resistance and antiretroviral use are added to the adjustment for people living with HIV. * The ORs for the Ralph/Timika score and percent of lung involved in disease are per 10-point increase.

| Feature | Unadjusted OR<br>(95% CI) | P-value<br>(OR) | Adjusted OR<br>(95% CI) | P-value<br>(aOR) |
| --- | --- | --- | --- | --- |
| **People living without HIV + rifampin-susceptible TB (Rif-S1, n = 566)** |  |  |  |  |
| **Timika\*** | 1.12 (1.06, 1.19) | <0.001 | 1.07 (1.00, 1.14) | 0.066 |
| **PLI\*** | 1.16 (1.08, 1.26) | <0.001 | 1.11 (1.01, 1.22) | 0.025 |
| **Cavities (vs. none)** |  |  |  |  |
| Presence | 1.69 (1.06, 2.69) | 0.027 | 1.12 (0.65, 1.91) | 0.688 |
| Small | 1.45 (0.87, 2.41) | 0.150 | 1.02 (0.57, 1.81) | 0.954 |
| Medium | 1.69 (0.96, 2.99) | 0.071 | 1.35 (0.72, 2.52) | 0.353 |
| Large | 1.21 (0.56, 2.58) | 0.628 | 0.94 (0.42, 2.10) | 0.875 |
| Multiple | 1.28 (0.70, 2.32) | 0.425 | 0.89 (0.46, 1.72) | 0.733 |
| **Lymphadenopathy** | 0.75 (0.43, 1.29) | 0.293 | 0.74 (0.41, 1.35) | 0.331 |
| **Nodules (presence)** | 2.43 (1.14, 5.21) | 0.022 | 2.05 (0.91, 4.62) | 0.082 |
| **Pleural effusion** | 0.93 (0.51, 1.70) | 0.816 | 1.06 (0.54, 2.06) | 0.872 |
| **People living without HIV + rifampin-resistant TB (Rif-R1, n = 1,056)** |  |  |  |  |
| **Timika\*** | 1.19 (1.14, 1.24) | <0.001 | 1.14 (1.08, 1.20) | <0.001 |
| **PLI\*** | 1.28 (1.21, 1.36) | <0.001 | 1.21 (1.13, 1.30) | <0.001 |
| **Cavities (vs. none)** |  |  |  |  |
| Presence | 2.01 (1.48, 2.74) | <0.001 | 1.55 (1.09, 2.21) | 0.015 |
| Small | 1.72 (1.26, 2.34) | 0.001 | 1.41 (1.00, 2.01) | 0.053 |
| Medium | 1.94 (1.33, 2.83) | 0.001 | 1.65 (1.08, 2.52) | 0.020 |
| Large | 3.47 (2.22, 5.41) | <0.001 | 3.21 (1.93, 5.33) | <0.001 |
| Multiple | 2.85 (1.99, 4.09) | <0.001 | 2.49 (1.65, 3.75) | <0.001 |
| **Lymphadenopathy** | 1.42 (0.97, 2.08) | 0.068 | 1.36 (0.89, 2.08) | 0.154 |
| **Nodules (presence)** | 1.21 (0.75, 1.95) | 0.430 | 0.94 (0.56, 1.58) | 0.814 |
| **Pleural effusion** | 1.36 (0.87, 2.13) | 0.180 | 1.15 (0.70, 1.91) | 0.580 |
| **People living with HIV + any TB (HIV, n = 372)** |  |  |  |  |
| **Timika\*** | 1.09 (1.03, 1.16) | 0.003 | 1.08 (1.01, 1.16) | 0.016 |
| **PLI\*** | 1.10 (1.02, 1.20) | 0.014 | 1.11 (1.01, 1.21) | 0.027 |
| **Cavities (vs. none)** |  |  |  |  |
| Presence | 1.77 (1.12, 2.79) | 0.014 | 1.55 (0.92, 2.63) | 0.103 |
| Small | 1.57 (0.97, 2.53) | 0.064 | 1.42 (0.82, 2.46) | 0.216 |
| Medium | 3.23 (1.35, 7.76) | 0.009 | 2.69 (0.99, 7.33) | 0.053 |
| Large | 2.55 (0.6, 10.82) | 0.205 | 1.72 (0.30, 9.93) | 0.546 |
| Multiple | 2.21 (1.14, 4.30) | 0.019 | 1.80 (0.84, 3.83) | 0.129 |
| **Lymphadenopathy** | 0.93 (0.59, 1.48) | 0.769 | 1.08 (0.64, 1.81) | 0.778 |
| **Nodules (presence)** | 1.32 (0.65, 2.67) | 0.439 | 0.84 (0.38, 1.86) | 0.661 |
| **Pleural effusion** | 1.12 (0.66, 1.89) | 0.674 | 1.02 (0.56, 1.86) | 0.951 |

### PLI improves treatment outcome prediction accuracy when combined with clinical variables

We combined the Rif-S1 and Rif-R1 groups (n = 1,622) to boost statistical power. We trained logistic regression models on 75% of the data (n = 1,216) and assessed their generalizability to the remaining 25% (n = 406). We evaluated seven single-variable radiological models (**Fig. S3A, D**). PLI and Timika had the highest accuracy [AUC _(PLI)_ 0.656 (0.595, 0.717)], and the former performed significantly better than cavitation [ΔAUC _(PLI-Cavities (size))_ 0.065 (0.000, 0.130), p-value 0.034]. The addition of cavitary disease to PLI did not improve accuracy [ΔAUC _(PLI–PLI+Cavities (y/n))_ -0.001 (-0.023, 0.021), p-value 0.590] indicating that the predictive accuracy of Timika is derived predominantly from the PLI component (**Fig. S3B, D**). Given that smear grade (SG) was the only clinical variable associated with outcome across the three subgroups, and the recognition of the role of the basic demographics of age and sex from prior literature, we tested the addition of PLI to a sex+age and a sex+age+SG models respectively. For the former the addition of PLI significantly improved accuracy [ΔAUC _(sex+age+PLI–sex+age)_ 0.052 (0.011, 0.093), p-value 0.012] but did not reach the performance of the *complete* 13 non-radiological variable model (**Fig. S3C, D**). The change in AUC resulting from addition of PLI was similar in magnitude by resistance group but was only statistically significant for the Rif-R1 group (**Table S4**). We found similar trends in improvement of prediction with the addition of PLI to sex+age+SG, but this improvement did not reach statistical significance in the training:validation dataset [AUC _(sex+age+SG)_ 0.683 (0.630, 0.736) vs. AUC _(sex+age+SG+PLI)_ 0.714 (0.660, 0.768), p-value 0.057](**Fig. S4**). We repeated the analysis with people with HIV and observed similar improvements in prediction when PLI was added to the sex+age model but the increases were not statistically significant [ΔAUC _(sex+age+PLI–sex+age)_ 0.050 (-0.029, 0.129), p-value 0.080] (**Fig. S5**).

### PLI improves prediction accuracy in independent data

We used a chronologically independent dataset of patients without HIV (Rif-S2: 2020-2023, Rif-R2: 2021-2023) to validate model accuracy (n = 815). This sample had a similar distribution of sex, prior TB, and rifampicin resistance as the training-validation data (**Table 1**) but was skewed geographically (85% from Ukraine) (**Fig. S1C**), and had a higher frequency of anemia, other comorbidities, smoking, alcohol use, and high smear grade. On this independent data, we observed a similar increase in model accuracy with the addition of Timika or PLI to sex+age as we observed in the training-validation set [ΔAUC _(sex+age+PLI–sex+age)_ 0.054 (0.028, 0.080), p-value <0.001] (**Fig. 2**). The addition of PLI to a model with sex+age+SG also improved prediction accuracy in the test dataset [ΔAUC _(sex+age+SG+PLI–sex+age+SG)_ by + 0.028 (0.006, 0.050), p-value 0.004] (**Fig. S4**).

**Figure 2.**
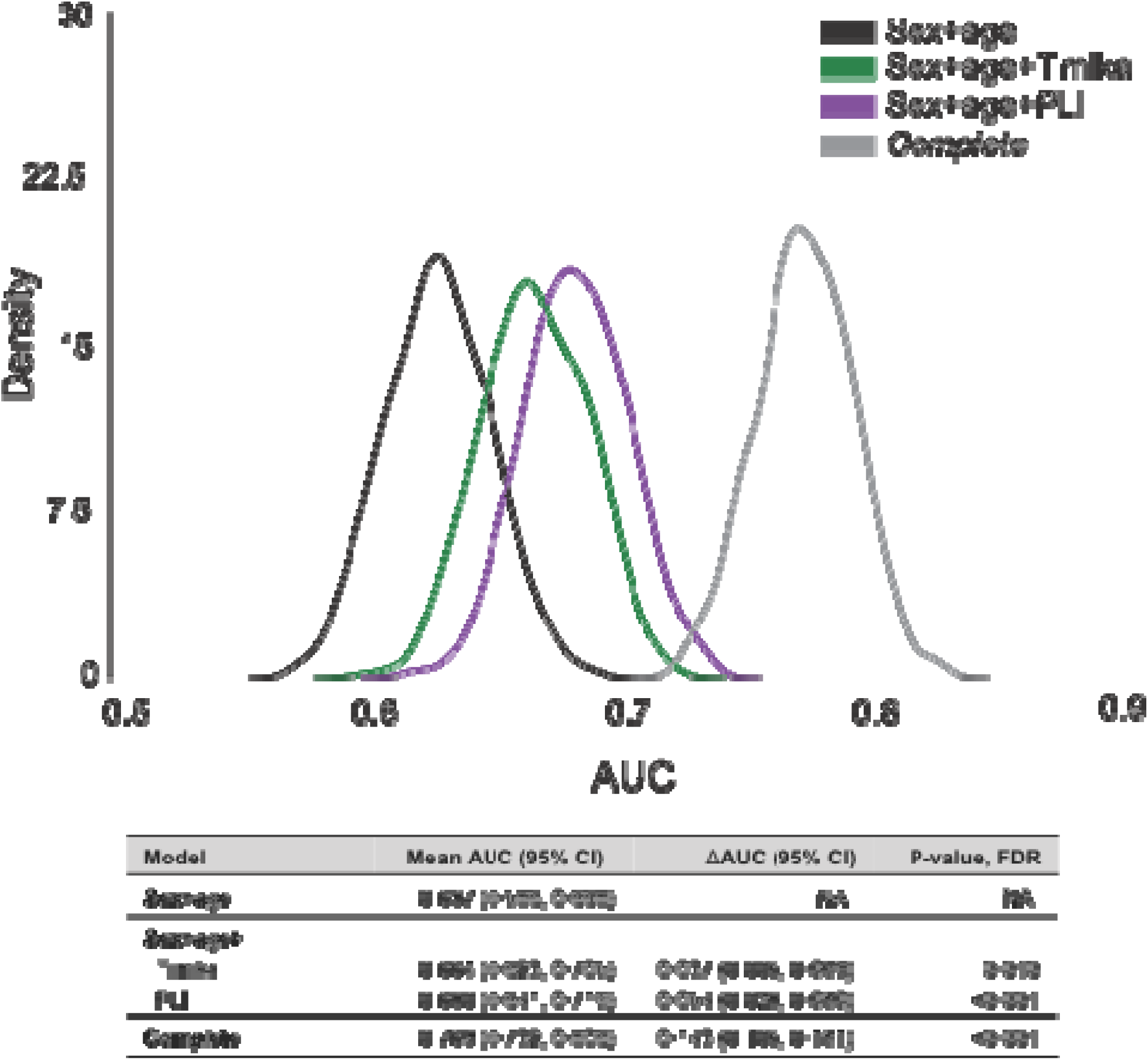
Prediction of unfavorable outcomes for radiological features (validation). We trained the logistic regression models on Rif-S1 + Rif-R1 (n = 1,622) and predicted outcomes on Rif-S2 + Rif-R2 (n = 815). We used sampling with replacement (1,000 iterations) on Rif-S2+Rif-R2 to generate a mean AUC and confidence intervals. The data represents the mean AUC of the 1,000 iterations and the 95% CI. At every iteration, we computed the difference between model AUCs (ΔAUC), and the number of observed differences that were ≤ 0 were divided by the total number of observations to assess statistical significance using a one-tailed empirical p-value approach [P-value = (#ΔAUC) ≤0/1,000]. We corrected for multiple hypothesis testing by controlling the Benjamini-Hochberg false discovery rate to <0.05. ΔAUC and P-values were computed by comparing each model to the sex+age model. The KDE plots are visual representations of the mean AUC and 95% confidence interval for the reduced model +/- PLI or Timika.

### Impact of radiology on the stratification of risk

To understand the clinical implications of using radiology for baseline TB risk assessment, we tuned the probability threshold defining high vs. low risk to maintain sensitivity at >98% for predicting unfavorable outcome (**Methods**, **Fig. 3A**). This allows for a scenario in which the risk assessment focuses on ruling *out* unfavorable outcomes. We then tested the optimal threshold for each model on the independent dataset. Sex+age+SG+PLI specificity increases to 14.9% from 11.6% vs. sex+age+SG, approaching the specificity of the *complete* model (15.1%), with comparable sensitivity (sex+age+SG 98.6%, sex+age+SG+PLI 98.6%, complete 99.1%) (**Fig. 3B**). In absolute numbers, the addition of PLI to sex+age+SG increases the size of the low-risk group from 73 (9.0% of total, n = 3 unfavorable outcome) to 93 (11.4% of total, n = 3 unfavorable outcome) in the independent data (n = 815) (**Table S5**).

**Figure 3.**
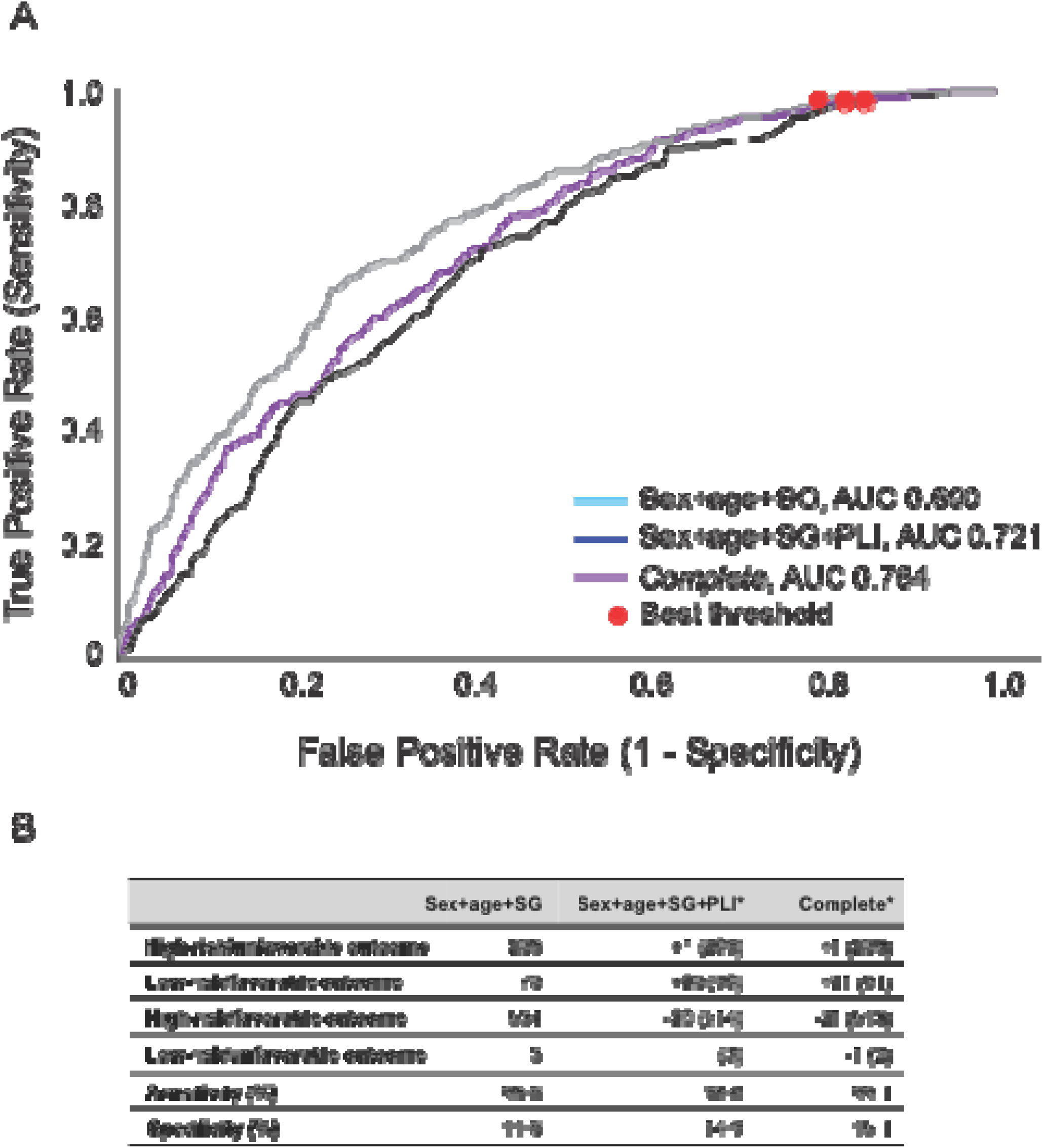
Finding the optimal model threshold to separate low- and high-risk groups. (A) ROC for sex+age+SG (black), sex+age+SG+PLI (purple) and complete (gray) logistic regression models for unfavorable outcomes. Filled red circles represent the optimal threshold for a sensitivity ≥ 0.98 with the maximum geometric mean for sensitivity and specificity. (B) Breakdown of statistics for each model’s optimal threshold when applied on the training (n = 1,622) and validation (n = 815) datasets. * Values are represented as the difference from values in the sex+age column.

### Optimal threshold of PLI and Timika score dichotomization

A PLI cutoff of 50% was previously suggested as a predictor of unfavorable treatment outcome(14, 15). We studied the optimal threshold on PLI using Monte Carlo cross-validation. Using the training-validation set of people without HIV (n = 1,622), we identified the optimal threshold for PLI at 25%, and for Timika at 56/140 (**Fig. 4A**) to maximize the geometric mean of sensitivity and specificity. PLI at 25% had higher sensitivity compared with PLI at 50% (sensitivity-specificity of 59.7-65.3 vs. 25.6-88.9 respectively) and increased the size of the high-severity group by 171% (high-risk: 333 vs. 121 out of total n=815 respectively) (**Fig. 4B**).

**Figure 4.**
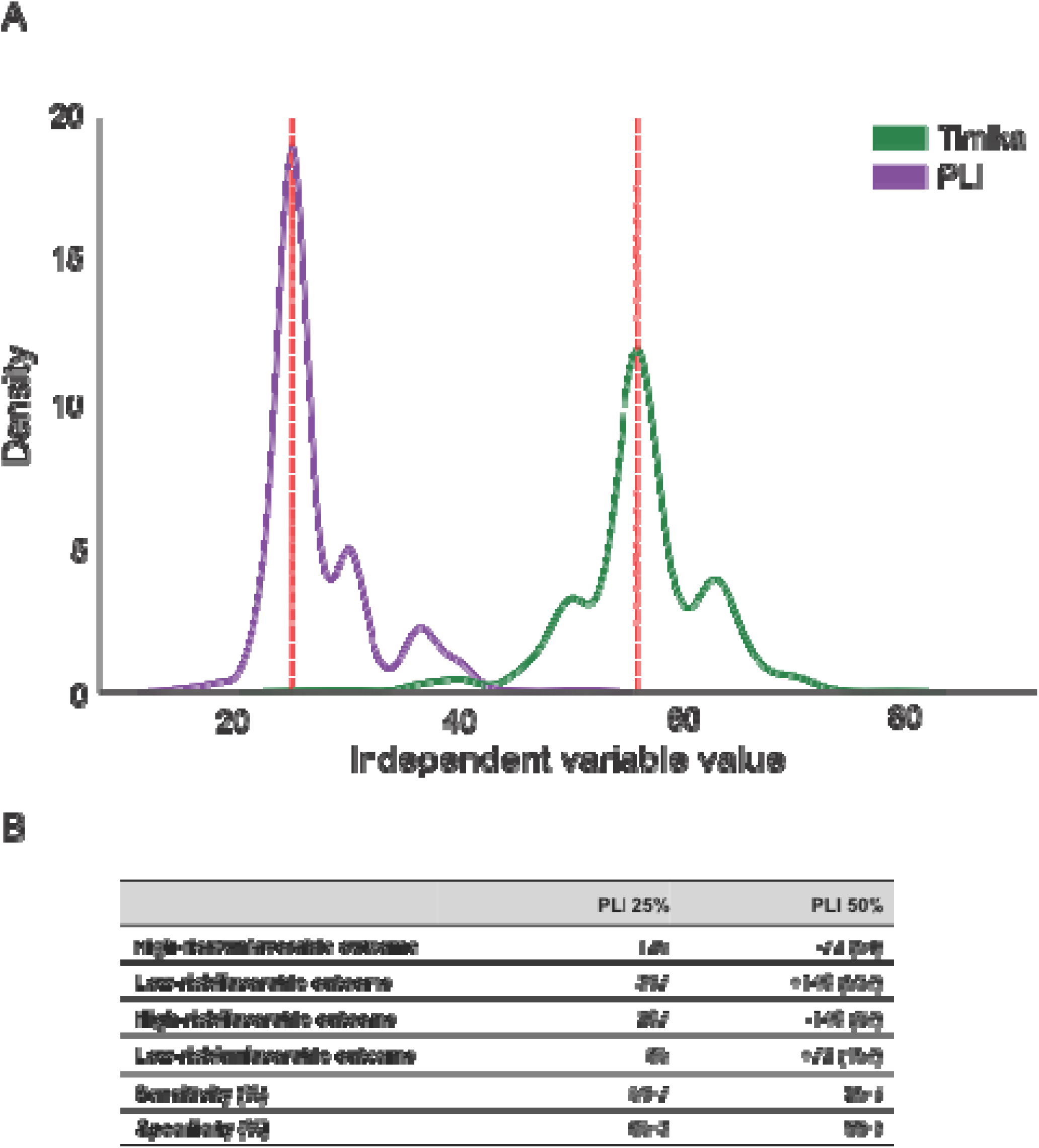
Optimal threshold for binarizing PLI. We built PLI-only and Timika-only logistic regression models and estimated the prediction accuracy for unfavorable outcomes using Rif-S1 + Rif-R1 (n = 1,622). We used a Monte Carlo cross-validation approach with 1000 iterations of resampling (75:25). (A) The KDE plots are visual representations of the mean AUC and 95% confidence interval. Optimal thresholds for raw values of PLI and Timika score based on optimal trade-off between sensitivity and specificity (maximal geometric mean) for the individual radiological feature models for PLI or Timika (red dashed line = median for optimal threshold). (B) Breakdown of statistics for thresholding at PLI 25% vs. 50% when models are trained on training-validation dataset (Rif-S1+Rif-R1, n = 1,622) and tested on test data (Rif-S2+Rif-R2, n=815).

### PLI in external severity scores of unfavorable outcomes

We benchmarked TB severity scores: A5414/SPECTRA-TB *(protocol in development)* and endTB-Q(6) (ClinicalTrials.gov NCT03896685) as they both incorporate radiological findings and are currently being investigated as a guide for shortening TB treatment in two randomized clinical trials. We assessed the accuracy of these scores in predicting treatment outcome in real-world TB care settings and, assessed how the real-world accuracy of these scores changes with the use of PLI 25% or PLI instead of cavitation. A5414/SPECTRA-TB scores severity in drug-susceptible TB based on data from S31/A5349(7) (ClinicalTrials.gov <u>NCT02410772</u>) assessing a four-month rifapentine-containing treatment regimen for drug-susceptible TB, and includes extent of disease at PLI ≥ 50% (SPECTRA50, *manuscript under review*). We compared this model to a modified SPECTRA25 model (PLI ≥ 25%), sex+age+SG+PLI and *complete*+PLI trained on Rif-S1 (n=566) and tested on the pooled Rif-S data (n=851) (**Supplementary Methods**). There was no statistically significant difference between the AUCs for SPECTRA50, SPECTRA25, and sex+age+SG+PLI, but the modified SPECTRA25 score had a slightly higher mean AUC than the SPECTRA50 score (AUC 0.689 and 0.678, respectively) (**Fig. S6A-B**). endTB-Q uses smear grade and cavity presence to predict severity in drug-resistant TB(6). We compared a logistic regression model based on endTB-Q to a modified endTB-Q_PLI (replacing cavity presence with PLI (0-100)), sex+age+SG+PLI and *complete*+PLI (**Supplementary Methods**) trained on Rif-R1 (n = 1,056) and tested on Rif-R (n=1,586). Both endTB-Q_PLI [ΔAUC _(endTB-Q_PLI–endTB-Q)_ 0.063 (0.035, 0.091), p-value <0.001] and sex+age+SG+PLI [ΔAUC _(sex+age+SG+PLI–endTB-Q)_ 0.124 (0.091, 0.157), p-value <0.001] performed better than endTB-Q [AUC 0.602 (0.571, 0.633)] (**Fig. S6C-E**)

### Artificial intelligence to accurately predict PLI and Timika

Computer-assisted diagnosis (CAD) uses artificial intelligence (AI) to automate TB diagnosis from CXR and has gained rapid clinical adoption globally. CAD is trained to classify TB disease as present or absent(20, 21) and has recently been suggested for use in disease severity (22). We trained an AI model to classify TB disease severity from CXR focusing on PLI (both continuous and binarized at 25%) and Timika (continuous and binarized at 55). From TB-Portals, 5,261/7,213 chest X-ray DICOM images passed quality control for use in AI (**Supplementary Methods**). Of these, 2,893 were Rif-S and 2,368 were Rif-R. The ensemble CNN model (DenseNet121-res224-all) had the highest accuracy for predicting PLI and Timika score, independently and jointly for Rif-S and Rif-R data subsets [test MAE 11.7 (95%CI 10.6-12.8) and 15.8 (95%CI 14.6-17.0) respectively; test AUC 0.86 (95%CI 0.82-0.88) and 0.78 (95%CI 0.73-0.83) respectively] (**Table S6**).

## Discussion

We show that among ten CXR findings in pulmonary TB, PLI is most consistently associated with unfavorable treatment outcome. Cavitation improves model fit for the rifampicin-resistant group, but does not improve the prediction of unfavorable outcome when added to PLI. PLI improves prediction of unfavorable outcome over demographics with and without smear grade. The use of PLI increases the size of low-risk class of patients compared to demographics alone and may be helpful to increase the number of patients successfully treated with a less intense or shorter regimen, when such regimens become available (ex. ClinicalTrials.gov NCT02410772)(6, 7, 15, 23).

Our study shows that CXR findings are not sufficiently accurate as standalone predictors of outcome and emphasizes the importance of combining radiology with baseline demographics and readily available microbiological metrics to achieve acceptable accuracy (AUC >0.70). Based on our findings, we recommend using a model including sex, age, smear grade and PLI to provide informed predictions for poor outcomes in pulmonary TB. Other useful baseline variables in some subgroups include BMI, anemia, smoking and alcohol use however in the best-case scenario these combined only add ∼0.03 to the accuracy of the sex, age, smear grade and PLI model. We evaluated the A5414/SPECTRA-TB and endTB-Q clinical trial severity scores for predicting outcomes for real-world rifampicin-susceptible and rifampicin-resistant TB, respectively. The definitions and ascertainment of unfavorable outcomes differ between clinical trial and real-world settings. In the former, recurrence of disease and/or complex composite outcomes and adherence are typically captured but not in the latter. Despite these differences, the AUCs for outcome prediction are comparable across data from these two settings (Yu A et al, unpublished). The performance of A5414/SPECTRA-TB may be improved if PLI at 25% is used to replace PLI at 50% and that of endTB-Q improved if PLI is used to replace cavitation and used as a continuous value. Validation on external data and specifically on data from clinical trials of TB shortening is recommended to confirm these findings and better assess their implications.

Pulmonary cavitation in TB is thought to result from the necrosis and expansion of TB granulomas or diseased lung(24). After their formation during or in the recovery phase of active disease, cavities often persist in the lung chronically and/or lifelong(3). The presence and size of cavitation have been previously linked to unfavorable treatment outcomes, and used to describe severity in clinical trial settings(4, 5, 7, 12). PLI describes the proportion of opacified lung parenchyma, a process expected to start earlier than cavitation and that subsides after recovery and cure(11, 13). We observe a stronger association for baseline PLI and outcome than for cavitation and outcome, and we identify no added predictive role of cavitation over PLI alone. We speculate that previous associations of cavitation with unfavorable outcomes in drug-susceptible disease may have been related to a correlation between cavitation and PLI in the subacute setting, *i.e.* patients with more extensive parenchymal disease may be more likely to progress to cavitation. It is also possible that patients with a delayed presentation are more likely to have both extensive disease involvement and cavitation, as the latter takes more time to develop.

People living with HIV can have more subtle TB findings on CXR than people without HIV(25). This is believed to be due to ineffective recruitment of immune cells to the site of disease. We observed lower prevalence of cavitary disease and lower Timika scores in the HIV group compared to the non-HIV groups. PLI on the other hand is associated with unfavorable outcomes in the HIV group with a similar effect size to that observed for the non-HIV group. This suggests that PLI is an appropriate universal measure of radiological TB severity.

As digital CXR technology is now readily available in most TB treatment settings, the use of AI can automate interpretation, potentially improve accuracy and reduce inter-reader variability. We were able to accurately automate PLI thresholding at 25% and further work should validate these models prospectively across different geographic settings and directly in risk stratification.

Our study had several limitations including its retrospective nature that poses a risk of unmeasured confounding. Nevertheless, one strength is that we synthesized data across several cohorts lessening the impact of bias or mismeasurement from an individual cohort on the overall conclusions. We note that the *complete* models of outcome focused on baseline variables. Although we did include an effective treatment variable to account for regimen composition and antibiotic resistance for improved isolation of effects of baseline variable, we were not able to account for adherence as this data is not collected by the programs and/or TB-Portals. The lack of control for adherence is common in retrospective modeling of TB treatment outcomes and affects most prior work on associating baseline CXR features and outcomes in TB. Another limitation is that human readers of the CXRs in the dataset including general practitioners and trained radiologist with inter-reader variability. Such limitations are expected in real-world data. We nevertheless provide one of largest evaluation of radiological predictors treatment outcomes in a multicohort setting to date. We acknowledge that the use of radiological features in severity scoring is dependent on the availability of imaging capture technology and this technology is not yet universally available in high TB prevalence settings. However, digitalized imaging has been increasingly adopted as it becomes less expensive, and the use of automation including CAD has further reduced costs and increased access.

This work builds on one prior analysis where PLI was found associate with unfavorable treatment outcomes in a smaller TB-Portals dataset (26). We innovate over prior work by demonstrating the added importance of clinical and microbiological variables over PLI for severity assessment in TB and by directly comparing PLI with cavitation and eight other radiological findings. We demonstrate for the first time the universality of PLI across key TB subgroups including drug susceptible, drug resistant and HIV co-infection. We evaluate the implications of using PLI and its optimal threshold in severity scores currently used in clinical trials, and lastly developed new and accurate AI models for automating PLI quantitative prediction and classification at ≥25% respectively. Our work guides the implementation of CXR data in severity assessment in research and clinical trials for shortening treatment. In future directions, we hypothesize that baseline PLI measurement may also prove helpful in predicting long term pulmonary sequelae of TB and recommend further study of this possibility.

## Materials and Methods

### Study population

We used the TB-Portals multi-cohort database curated by the National Institute of Allergy and Infectious Diseases (NIAID) (accessed on September 19^th^, 2023; see Data and Code Availability) **(Table S1)**. Inclusion criteria are summarized in **Figure 1**. **Table S2** summarizes the processing of radiological and treatment outcome variables. For each CXR, findings were coded by one clinician. Multiple clinicians provide these readings to avoid biasing the data to a single observer’s image reading practices.

### Data preprocessing

We split the data into three groups based on drug resistance and HIV coinfection: (a) without HIV + rifampicin-susceptible (Rif-S), (b) without HIV + rifampicin-resistant (Rif-R) and (c) with HIV (HIV). We split the Rif-S and Rif-R groups into training-validation (Rif-S1 and Rif-R1) and test (Rif-S2 and Rif-R2) datasets. To accomplish this, we split patient records based on date of registration for training-validation and testing. For Rif-S, we assigned all cases between 2008-2019 and 2021-2023 to the training-validation and test datasets, respectively, and randomly assigned the cases from 2020 using the train_test_split function from the SciKit-Learn(27) model selection toolkit (v1.1.3) to generate the final 1,622:815 (66:33) data split. For Rif-R, we assigned all cases between 2008-2020 and 2022-2023 to the training-validation and test datasets, respectively, and randomly assigned the cases from 2021 to create the final 1,622:815 data split. We used Rif-S1, Rif-R1 and HIV in parallel to build logistic regression models for association studies and model fit analyses. We used Rif-S1 and Rif-R1 with Monte-Carlo cross-validation (75:25) to test the predictive accuracy of logistic regression models. We used Rif-S2+Rif-R2 with resampling with replacement to validate the predictive accuracy findings.

### Outcome definition

Treatment outcomes were concordant with the 2013 WHO criteria.(28) Death (during the course of treatment), treatment failure (treatment termination or need for permanent regimen change of at least two drugs), and palliative care were considered unfavorable outcomes, while cure (treatment completion + bacteriological proof of conversion in three consecutive cultures at least 30 days apart) and treatment completion (treatment completion with no signs/symptoms of TB disease) were considered favorable outcomes (**Table S2**).

### Regression

We fit univariable and multivariable logistic regression models using the Logit tool from Statsmodels(29) Python library (v0.13.2) and the Newton-Raphson method. We built a *complete* non-radiological logistic regression model using available demographic, medical, social, microbiological, and treatment variables selecting variables based on their suspected or known association with treatment outcomes based on literature evidence(4, 16, 17). We built a *reduced* model composed of sex and age at onset of disease (sex+age) to model clinical scenarios in which other characteristics are unavailable, excluding features that are difficult (e.g. extrapulmonary disease) or impossible (e.g. effective treatment) to collect at baseline, and that may be missing (e.g. BMI or comorbidities). We tested a second version of the *reduced* model with smear grade (sex+age+SG) given its strong association with outcomes(4, 18). We used the same logistic regression approach for radiological models. We compared the goodness of fit of nested models using likelihood ratio tests (LRT) and performed hypothesis testing with a chisquared test, false discovery rate (FDR)-correcting P-values for multiple testing. For training-validation predictive accuracy, we conducted Monte Carlo cross-validation with 1,000 iterations. Specifically, for each iteration, we trained the logistic regression models on 75% of the data, and predicted on the remaining 25%. For testing, we trained the models on Rif-S1+Rif-R1, predicted on Rif-S2+Rif-R2 and applied resampling with replacement at 1,000 iterations to generate AUC distributions.

### Statistical analysis for testing model prediction on independent data

We used bootstrapping to generate AUC distributions for each tested logistic regression model to test predictive accuracy. For the training-validation dataset, the bootstrapping was in the form Monte Carlo cross-validation, splitting the dataset 75:25 at each iteration for 1,000 iterations. For the test dataset, the bootstrapping was done through resampling with replacement for 1,000 iterations. At every iteration, we computed the difference between model AUCs (ΔAUC), and the number of observed differences that were ≤ 0 were divided by the total number of observations to assess statistical significance using a one-tailed empirical p-value approach [p-value = (#ΔAUC) ≤0/1,000]. We corrected for multiple hypothesis testing by controlling the Benjamini-Hochberg false discovery rate to <0.05.

### Rule-out risk assessment

We tuned the logit probability thresholds on training-validation data to predict unfavorable outcome with maximal geometric mean sensitivity and specificity while sensitivity ≥ 0.98. We tested the specificity and true negative rate of this threshold and models on the test dataset.

### Optimal prediction threshold for PLI and Timika

Using Rif-S1 and Rif-R1, we built a logistic regression model using PLI or Timika dichotomized at every integer value between 5 and 95 with 1000x Monte-Carlo cross-validation (75:25). For each model, we calculated the sensitivity and specificity, and assigned the best threshold to the model that has the highest geometric mean of sensitivity and specificity. We then computed the median of the 1000 best thresholds for PLI and Timika to generate the final optimal threshold, and validated the model compared to 50% PLI on independent data.

### External severity scores

SPECTRA50 is a model that includes age, BMI, diabetes, smear grade and extent of disease on CXR (PLI ≥50% vs. <50%). This model is a version of the original model generated from the S31/A5349(7) clinical trial *(manuscript in review)* that was pretrained and modified to fit our data structure. endTB-Q(6) is a simple classfier that uses smear grade with cavity. We used pretrained models with pre-defined coefficients (SPECTRA50) or trained logistic regression models (endTB-Q, sex+age+SG+PLI, *complete*+PLI) on the training-validation dataset of interest (Rif-S1 or Rif-R1 separately). We also tested modified versions of these scores based on findings from our analysis (SPECTRA25 and endTB-Q_PLI). SPECTRA25 is identical to SPECTRA50 with extent of disease (≥25% vs. <25%) and endTBQ_PLI is smear grade with extent of disease (0-100). We tested performance on the full TB-Portals whole dataset divided by drug resistance (Rif-S or Rif-R separately). We compared these models based on their AUCs. We also tested endTB-Q and endTB-Q_PLI as simple classifiers to mimic the use of endTB-Q in the original manuscript (using PLI ≥25% as a cutoff for endTB-Q_PLI instead of cavity presence).

### Convolutional Neural Networks

We used pretrained CNN models from the TorchXRayVision(30) Python library (https://github.com/mlmed/torchxrayvision/) to perform patient-level regression and classification on quality-controlled CXR DICOM data from TB-Portals. We used DenseNet121-based regression on the whole lung in concordance with recent work demonstrating this approach as more effective than applying regression on a presegmented image(22). We split the dataset 80:10:10 across training-validation-test sets. We pretrained The CNNs on one or multiple benchmark datasets available through TorchXRayVision. We used the training dataset to fine-tune the pretrained CNN models on the prediction of PLI and Timika. We chose the best performing model from the validation set for generalizability assessment on the test set. We computed distributions for the AUC and mean absolute error (MAE) with bootstrapping. Further details are available in the Supplementary Materials.

## Data and materials availability

The TB-Portals dataset is made readily available to external collaborators through NIAID after signing a data use agreement. More information on the raw data is present in the TB-Portals website (https://tbportals.niaid.nih.gov). We base this papFer on all TB-Portals data available for download by <u>September 19th, 2023</u>. We wrote all the scripts for this project on Jupyter notebooks using Python 3.9.12 using the Harvard Medical School O2 cluster and made them available on GitHub at https://github.com/farhat-lab/tbp-severity-scoring.

## Data Availability

All data produced are available online at: https://tbportals.niaid.nih.gov/

## Acknowledgments

We express our gratitude to Pranav Rajpurkar, PhD, and Emma Chen, MS, for their valuable advice on AI model selection and implementation relevant to chest X-ray datasets.

## Author Contributions

Conceptualization and design: MG, MRF

Methodology: MG, RS, YE, MRF

Model design and modification: MG, MRF, AYX, EY, LH, GEV, RMS

Data acquisition: AR, AG, DH, ZY, GR

Data analysis: MG, RS, YE, MRF

Result interpretation: MG, RS, YE, MRF, AYX, EY, LH, GEV, RMS, AR, AG, DH, ZY, GR, KRJ

Visualization: MG, MRF

Supervision: MRF, KRJ

Writing – original draft: MG, RS, YE, MRF

Writing – review & editing: AYX, EY, LH, GEV, RMS, AR, AG, DH, ZY, GR, KRJ

## Classification

Biological sciences: medical sciences

## Supplementary Materials for “Tuberculosis disease severity assessment using clinical variables and radiology enabled by artificial intelligence”

### Materials and Methods

#### Methods for ML-based prediction of radiological features

##### a. Cohort selection and image quality check

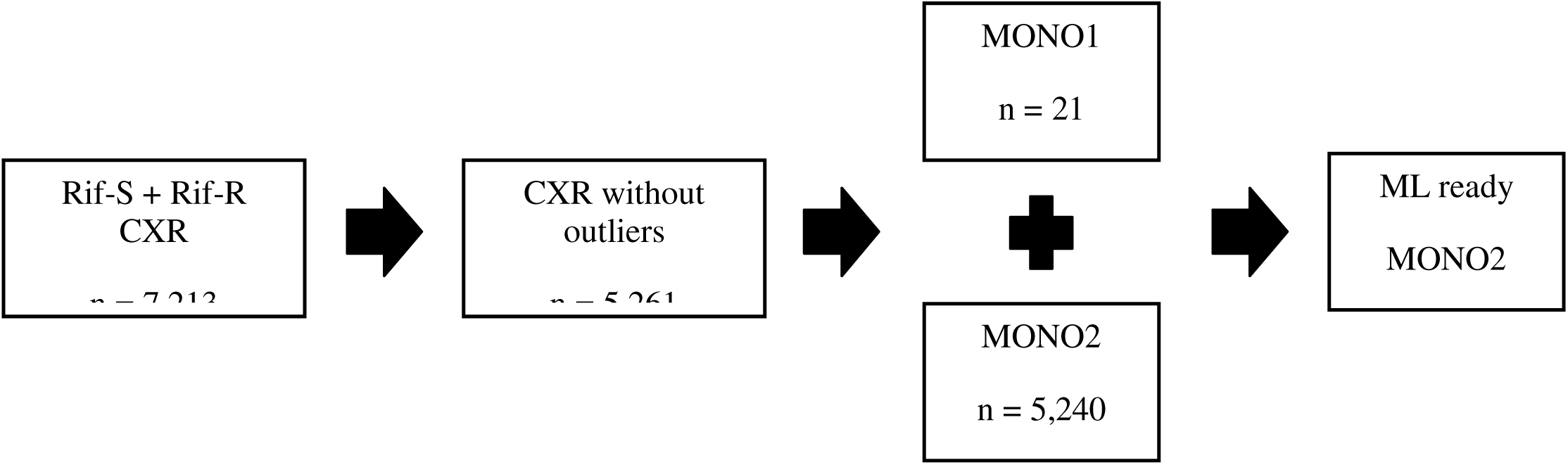

We included single-channel grayscale chest x-ray images and associated metadata in DICOM format. We used x-ray images that were closest to the treatment start date for two patient cohorts: rifampicin-sensitive (Rif-S, n=3,446) and rifampicin-resistant (Rif-R, n = 3,767). We only chose the patients that possessed complete set of values for % lung involvement and Timika score. We converted monochromatic medical images - Monochrome1 (MONO1) to Monochrome2 (MONO2) to harmonize the entire dataset to the same Photometric Interpretation attribute. We identified and removed outliers in the distribution of image quality using QC labels provided within TB-Portals. Outlier identification is based on a combination of both the programmatic results and the results of a human review of image data quality. The human review consisted of looking through the full set of public images and manually tagging images deemed unsuitable for machine learning.

##### b. Data pre-processing

We performed two classification tasks, including (a) percent of lung involved in disease (PLI, >25 or not) and (b) Timika score (>56 or not). For regression preprocessing, the raw PLI and Timika values were log-transformed to stabilize variance and then normalized using min-max scaling. Images were normalized using the TorchXRayVision library’s normalize function and resized to 224x224 pixels *(32).* The use of this library facilitated efficient implementation of deep learning models and ensured consistency in preprocessing steps across different datasets. The datasets were split 80:10:10 for training-validation:test. The test set contained unaltered data, to match the real-world data.

##### c. Model architecture and development

We used CNN models from TorchXRayVision *(31)* python library that were pretrained on chest x-ray image datasets, to perform patient-level regression as previously described *(22)* and classification tasks on PLI and Timika. We implemented two models: a first that was trained on the Stanford CheXpert dataset and another ensemble model trained on benchmark chest X-ray datasets including PadChest (University of Alicante), CheXpert (Stanford), MIMIC-CXR (MIT), Chest X-ray8 (NIH) and RSNA Pneumonia Challenge.

We first fit the model on the CXRs from TB-Portals originating from the two patient subgroups RIF-R (*n*□=□2,893) and RIF-S (n = 2,368) separately starting with representations from the CNN models that were pretrained on the available datasets in the TorchXRayVision library as above. The final model architecture is the pretrained DenseNet121 model without the final Classification layer. We repurposed the model to perform regression on the continuous values for PLI and Timika using Adam optimizer and calculated Mean Squared Error (MSE) Loss. Model performance was evaluated based on MSE and MAE through 100 epochs. We initially got similar performance on both the RIF-S and RIF-R subgroups (not shown), so we combined them and retrained the regression and classification models by calculating thresholds for PLI and Timika.

For the regression task, we utilized the DenseNet-121 architecture pretrained on chest X-ray datasets provided by the TorchXRayVision library *(33)*. The model’s final classifier layer was modified to predict a single continuous value. All layers except the final classifier layer were frozen to leverage the pretrained features effectively *(34)*. The Adam optimizer was employed with a learning rate of 0.0001 and Mean Squared Error (MSE) was chosen as the loss function *(34)*. The training process spanned 100 epochs with early stopping based on validation Mean Absolute Error (MAE) to prevent overfitting *(35)*.

For the classification task, the same DenseNet-121 architecture was employed but adjusted for binary classification by replacing the final classifier layer with a single output node and sigmoid activation function. The Binary Cross Entropy with Logits Loss (BCEWithLogitsLoss) was used as the loss function, and the Adam optimizer was employed with a learning rate of 0.001 *(36)*. Training also spanned 100 epochs with early stopping based on validation AUC to prevent overfitting.

### Supplementary Figures

**Supplementary Figure 1.**
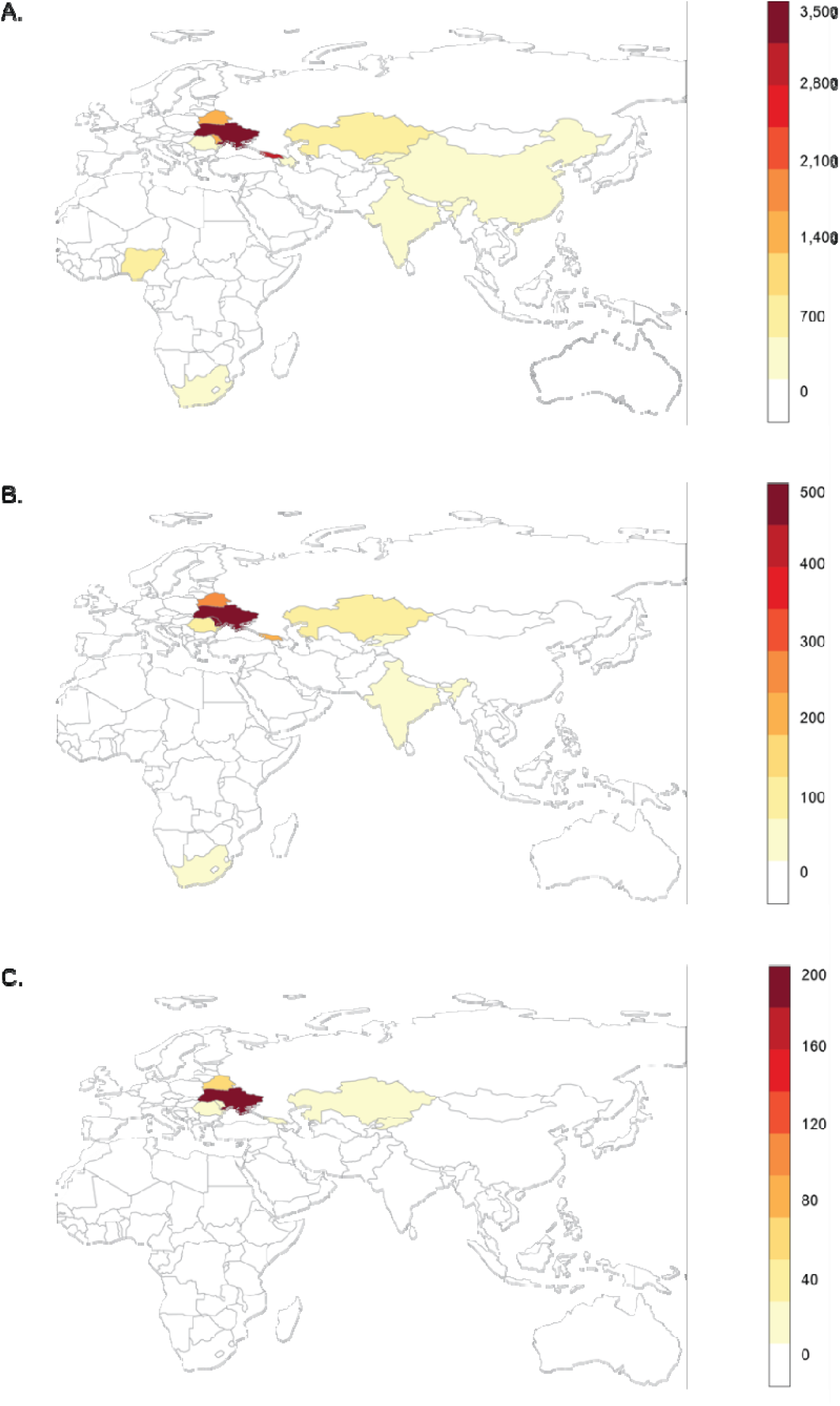
Geographic distribution of TB-Portals data. (A) Geographical distribution of all patients (first care episode) within the TB Portals database (n=11,067) with a scale ranging from 0-3,500 individuals. The dataset included patient cases from 13 countries: Ukraine, Georgia, Moldova, Belarus, Kazakhstan, Nigeria, Romania, Azerbaijan, South Africa, China, Kyrgyzstan, India, and Senegal. (B) Geographical distribution of cases included in the training-validation and HIV datasets (n=1,994) with a scale ranging from 0-500. (C) Geographical distribution of cases included in the test dataset (n=815) with a scale ranging from 0-200 individuals, respectively.

**Supplementary Figure 2.**
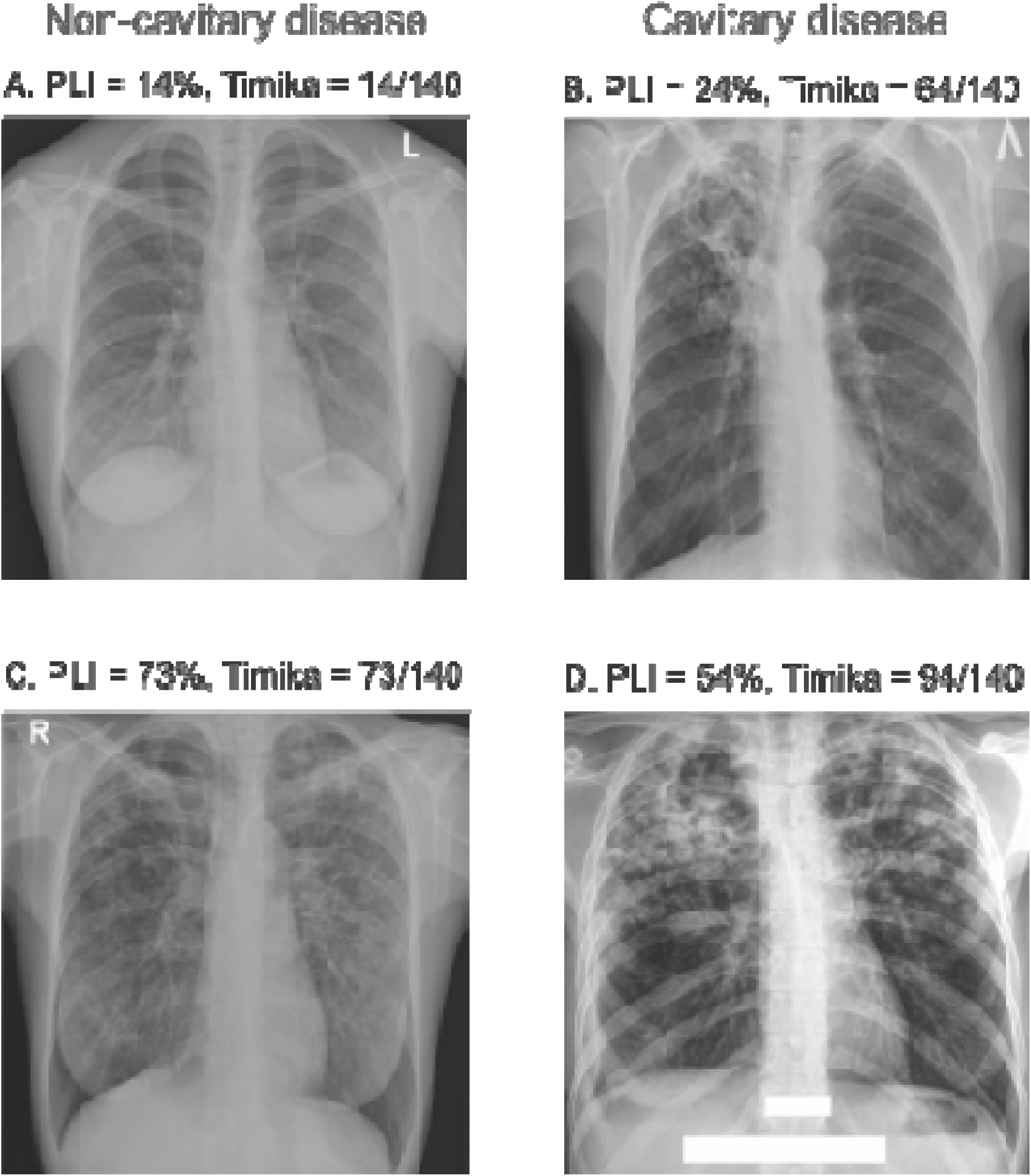
Examples of monochrome chest x-ray images present in the dataset. (A) low percent of lung involved in disease (PLI) and no cavitation, Timika score = PLI, (B) low PLI and cavitation, Timika score = PLI+40, (C) high PLI and no cavitation (Timika score = PLI) and (D) high PLI and cavitation, Timika score = PLI+40.

**Supplementary Figure 3.**
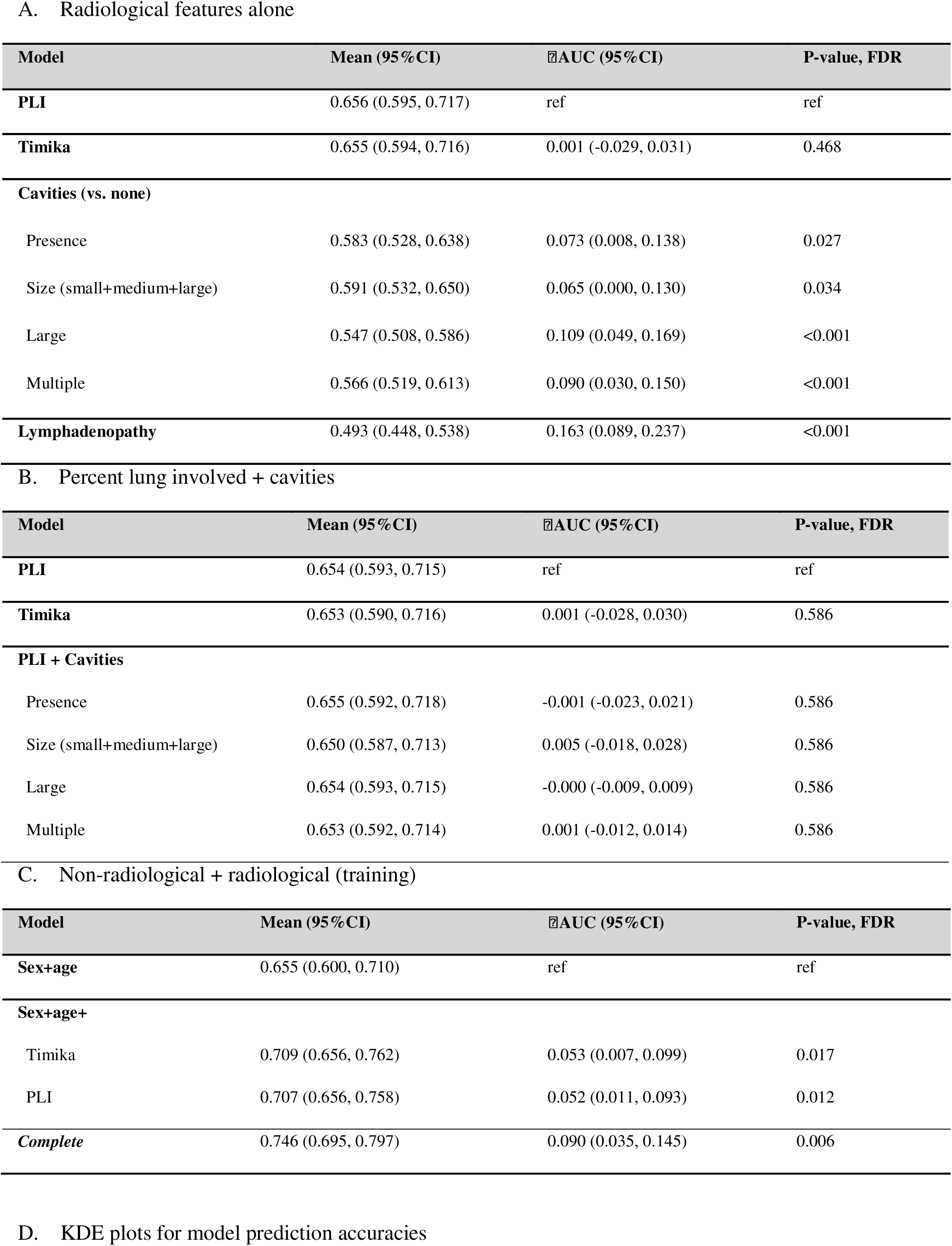
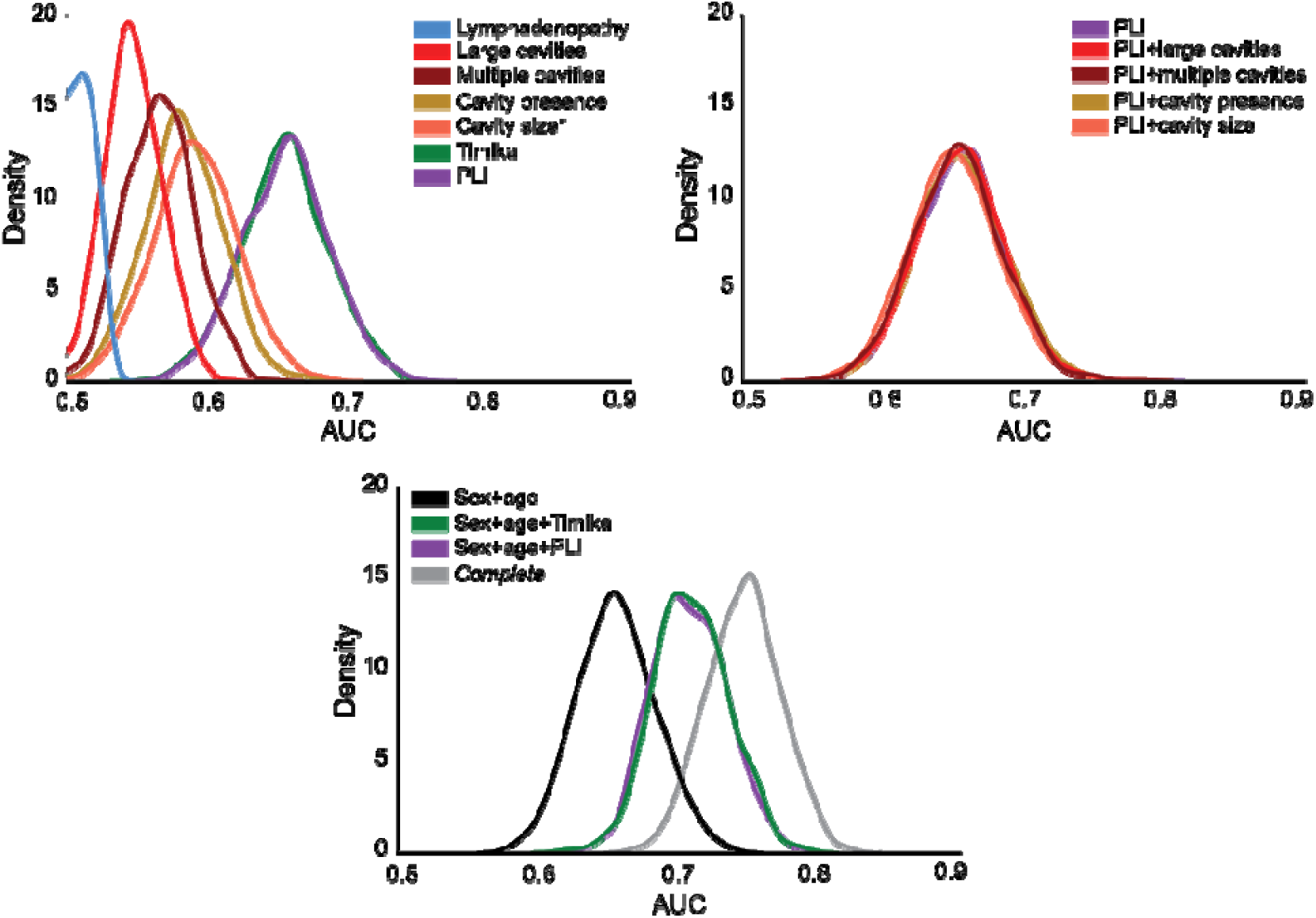
Prediction of unfavorable outcome using training data. We estimated the prediction accuracy using Rif-S1 + Rif-R1 (n = 1,622) for models built with (A) individual radiological features, (B) PLI + cavitary information and (C) non-radiological + radiological features. We used a Monte Carlo cross-validation approach with 1,000 iterations of resampling (75:25), trained on the 75% (n = 1,216) and tested of the 25% (n = 406) of each iteration. At every iteration, we computed the difference between model AUCs (ΔAUC), and the number of observed differences that were ≤ 0 were divided by the total number of observations to assess statistical significance using a one-tailed empirical p-value approach [p-value = (#ΔAUC) ≤0/1,000]. We corrected for multiple hypothesis testing by controlling the Benjamini-Hochberg false discovery rate to <0.05. In (A) and (B), IllAUC and FDR Pval were done compared to the PLI model, and in (C) IllAUC and FDR Pval were done compared to the sex+age model. (D) The KDE plots are visual representations of the mean AUC and 95% confidence interval for individual radiological features (top left), PLI +/- cavitary information (top right) and the reduced model +/- PLI or Timika (bottom).

**Supplementary Figure 4.**
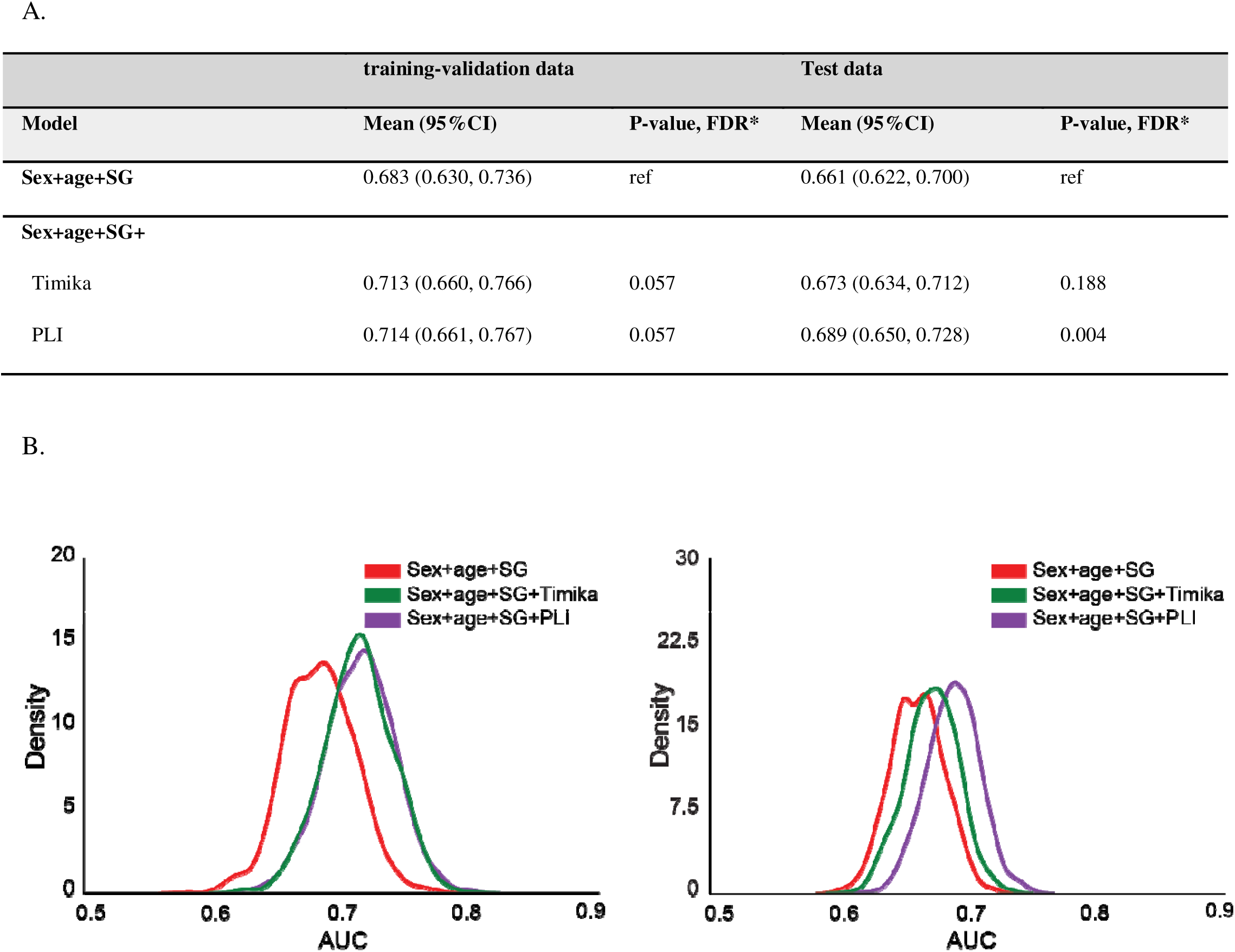
Prediction of unfavorable outcomes for radiological features with smear grade. (A) We estimated the prediction accuracy for models built with sex+age+smear grade +/- PLI or Timika. For the training-validation prediction accuracy assessment, we used a Monte Carlo cross-validation approach with 1000 iterations of resampling (75:25), trained on the 75% (n = 1,216) and validated of the 25% (n = 406) of each iteration. For the test dataset, we trained the logistic regression models on Rif-S1 + Rif-R1 (n = 1,622) and predicted outcomes on Rif-S2 + Rif-R2 (n = 815). We used sampling with replacement (1,000 iterations) to generate a mean AUC and confidence intervals. The data represents the mean AUC of the 1000 iterations and the 95% CI (mean +/- 1.96 x standard deviation). At every iteration, we computed the difference between model AUCs (ΔAUC), and the number of observed differences that were ≤ 0 were divided by the total number of observations to assess statistical significance using a one-tailed empirical p-value approach [p-value = (#ΔAUC) ≤0/1,000]. We corrected for multiple hypothesis testing by controlling the Benjamini-Hochberg false discovery rate to <0.05. IllAUC and FDR Pval were done compared to the sex+age+smear grade model. (B) The KDE plots are visual representations of the mean AUC and 95% confidence interval for the asex+age+smear grade model +/- PLI or Timika for the training-validation analysis (left) and the test analysis (right).

**Supplementary Figure 5.**
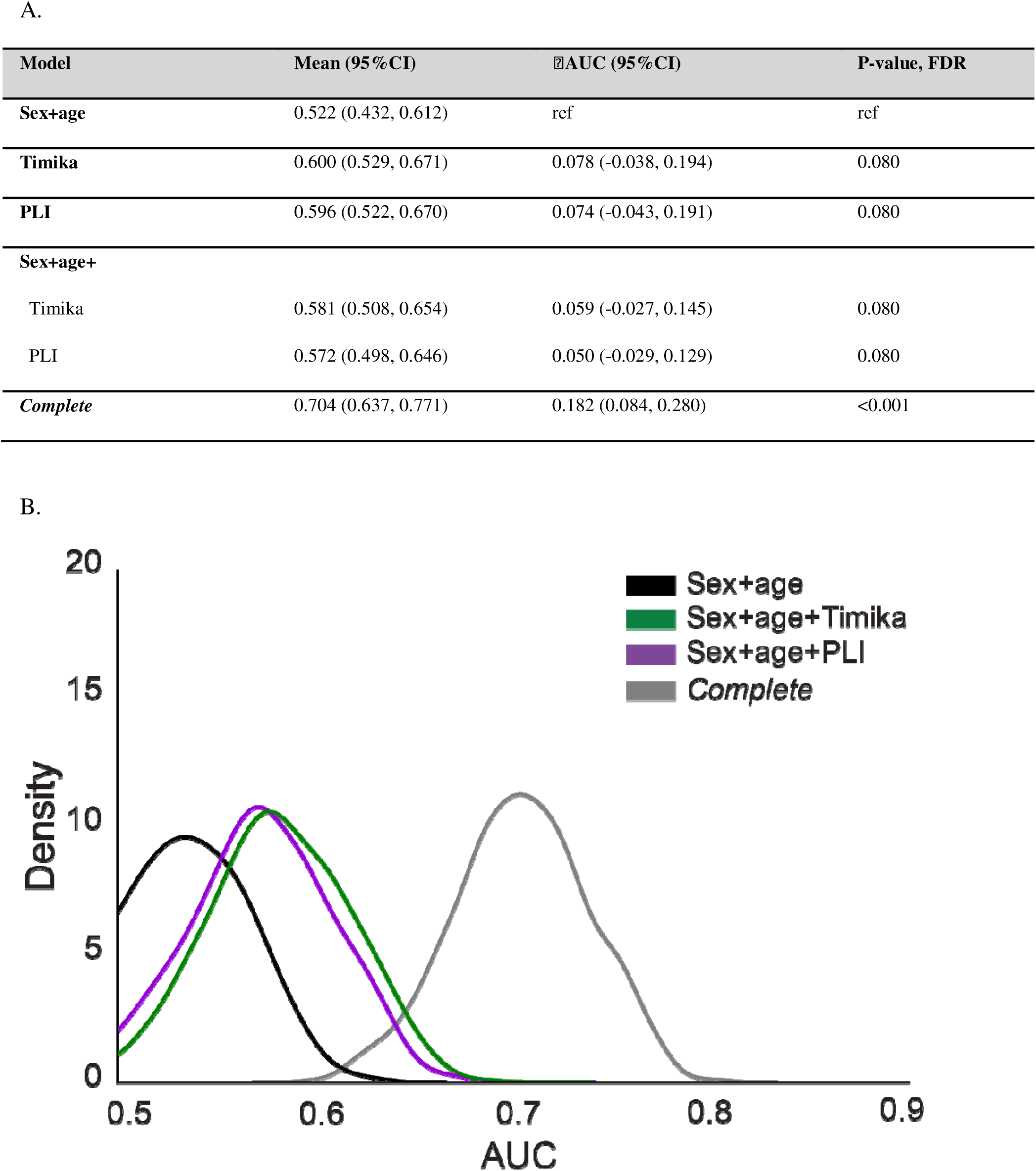
Prediction of unfavorable outcomes for radiological characteristics in people living with HIV. We estimated the prediction accuracy using HIV (n = 372). We used a Monte Carlo cross-validation approach with 1000 iterations of resampling (75:25), trained on the 75% (n = 223) and validated of the 25% (n = 149) of each iteration. (A) At every iteration, we computed the difference between model AUCs (ΔAUC). The data represents the mean AUC of the 1000 iterations and the 95% CI. The number of observed differences that were ≤ 0 were divided by the total number of observations to assess statistical significance using a one-tailed empirical p-value approach [P-value = (#ΔAUC) ≤0/1,000]. We corrected for multiple hypothesis testing by controlling the Benjamini-Hochberg false discovery rate to <0.05. IIAUC and the FDR-corrected P-value were done compared to the sex+age model. (B) The KDE plots are visual representations of the mean AUC and 95% confidence interval for the reduced model +/- PLI or Timika.

**Supplementary Figure 6.**
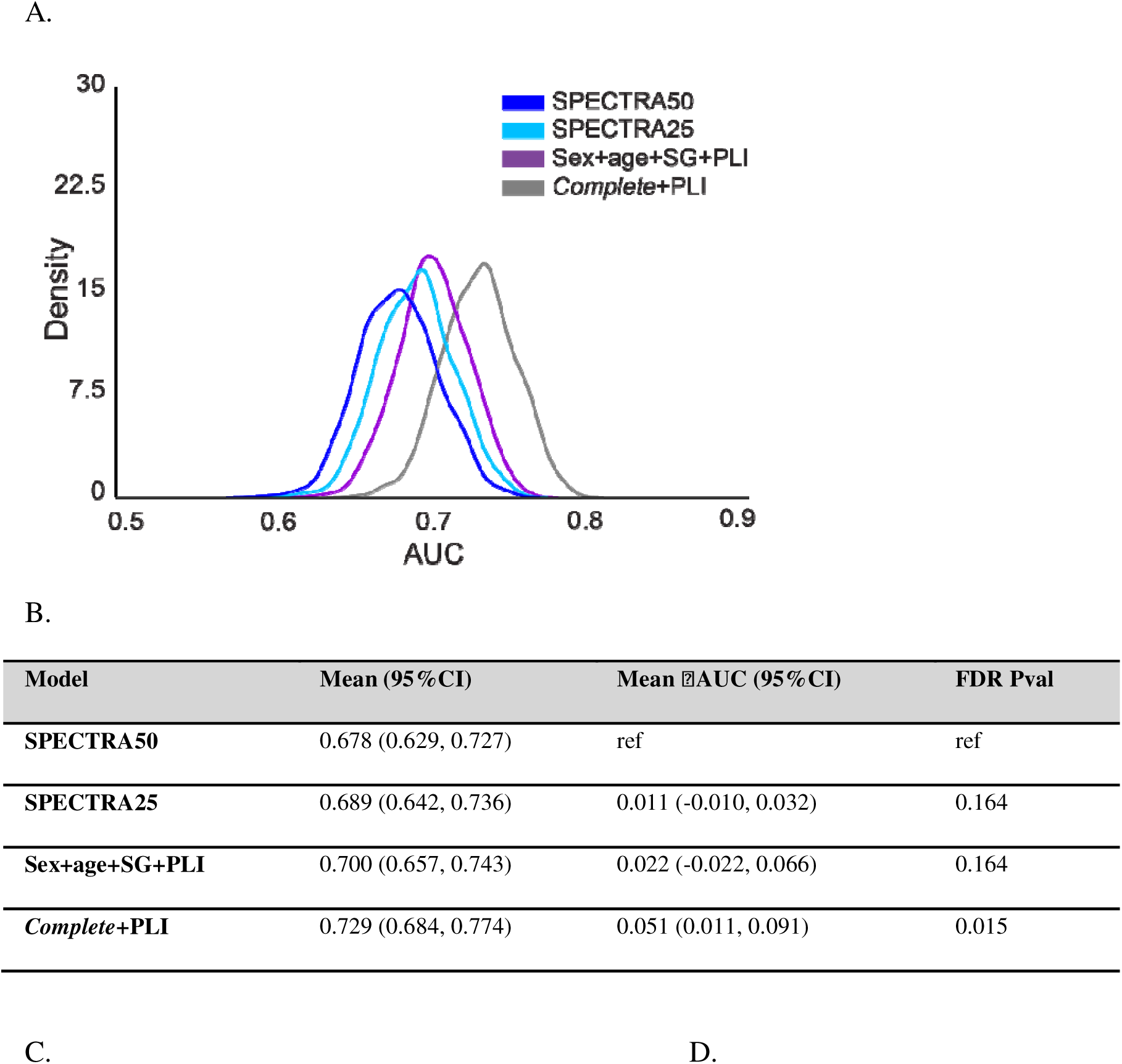
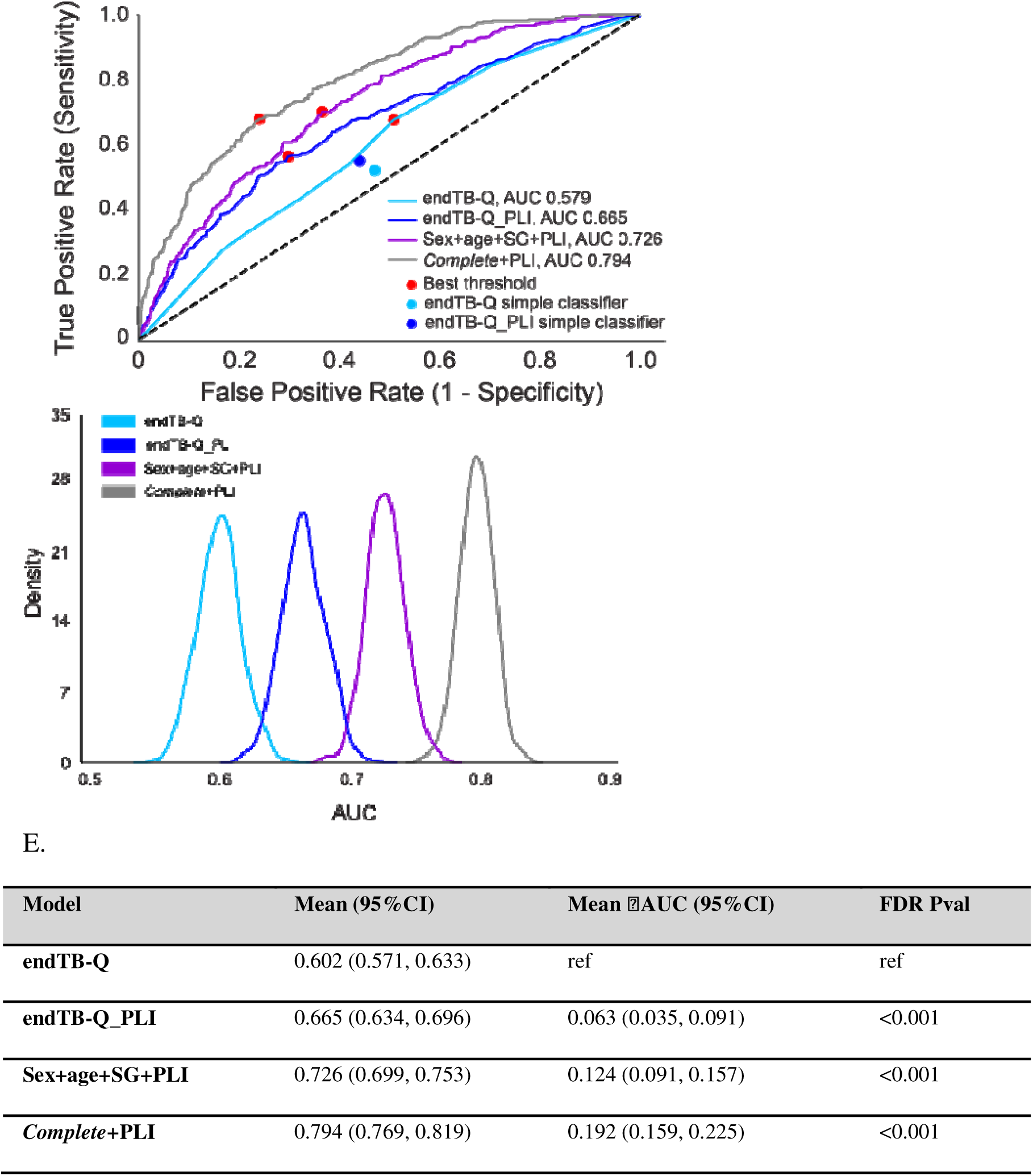
Testing SPECTRA with modifications for measuring severity in Rif-S and endTB-Q with modifications for measuring severity in Rif-R. We tested the performance of different severity models on (A, B) Rif-S and (C-E) Rif-R. (A, B) SPECTRA50 is a modified version of the SPECTRA model pretrained on data from the S31/A5349 4-month rifapentine regimen clinical trial and tested on Rif-S (n=851). SPECTRA25 uses the same coefficients as SPECTRA50 but lowers the threshold for PLI to 25% instead of 50%. Sex+age+SG+PLI and the complete models were trained on 2/3 of Rif-S (n=566) and tested on Rif-S (n=851). We used sampling with replacement (1,000 iterations) on Rif-S during the prediction task to generate a mean AUC and confidence intervals. The data represents the mean AUC of the 1,000 iterations and the 95% CI. At every iteration, we computed the difference between model AUCs (ΔAUC), and the data represents the mean ΔAUC of the 1,000 iterations and the 95% CI. the number of observed differences that were ≤ 0 were divided by the total number of observations to assess statistical significance using a one-tailed empirical p-value approach [p-value = (#ΔAUC) ≤0/1,000]. We corrected for multiple hypothesis testing by controlling the Benjamini-Hochberg false discovery rate to <0.05. IIAUC and FDR Pval compare each model to the SPECTRA50 model. (C-E) All models were trained on 2/3 of Rif-R (n=1,056) and tested on Rif-R (n=1,586). (C) Red dot: best threshold based on G-means of sensitivity and specificity. The endTB-Q simple classifier is the original classifier and the endTB-Q_PLI simple classifier is a modified classifier that includes a dichotomized extent of disease at PLI 25% instead of cavity presence. (D, E) We used sampling with replacement (1,000 iterations) on Rif-R during the prediction task to generate a mean AUC and confidence intervals. The data represents the mean AUC of the 1,000 iterations and the 95% CI. At every iteration, we computed the difference between model AUCs (ΔAUC), and the data represents the mean ΔAUC of the 1,000 iterations and the 95% CI. the number of observed differences that were ≤ 0 were divided by the total number of observations to assess statistical significance using a one-tailed empirical p-value approach [p-value = (#ΔAUC) ≤0/1,000]. We corrected for multiple hypothesis testing by controlling the Benjamini-Hochberg false discovery rate to <0.05. IIAUC and FDR Pval compare each model to the endTB-Q model.

### Supplementary Tables

**Supplementary Table 1.**
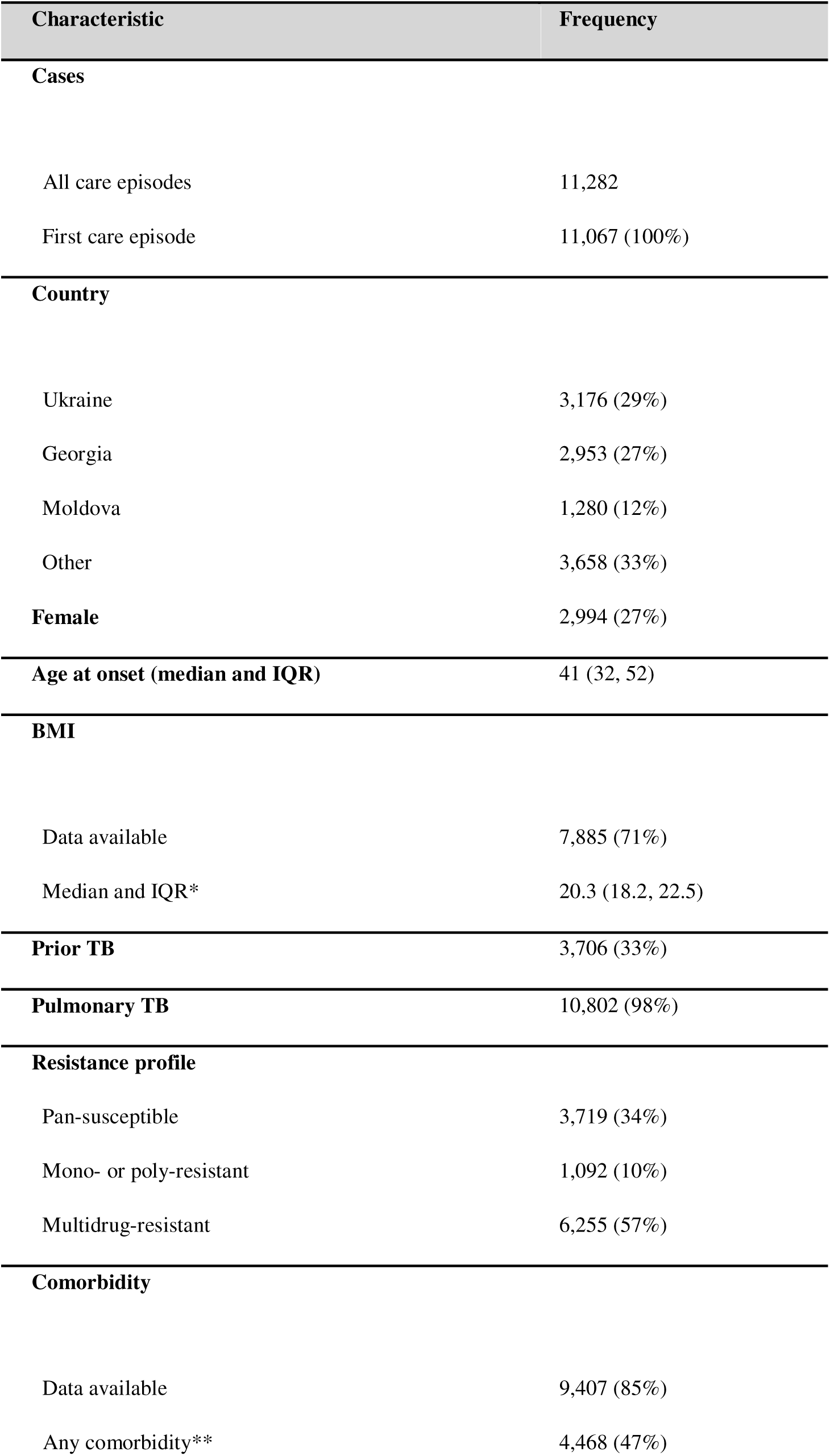
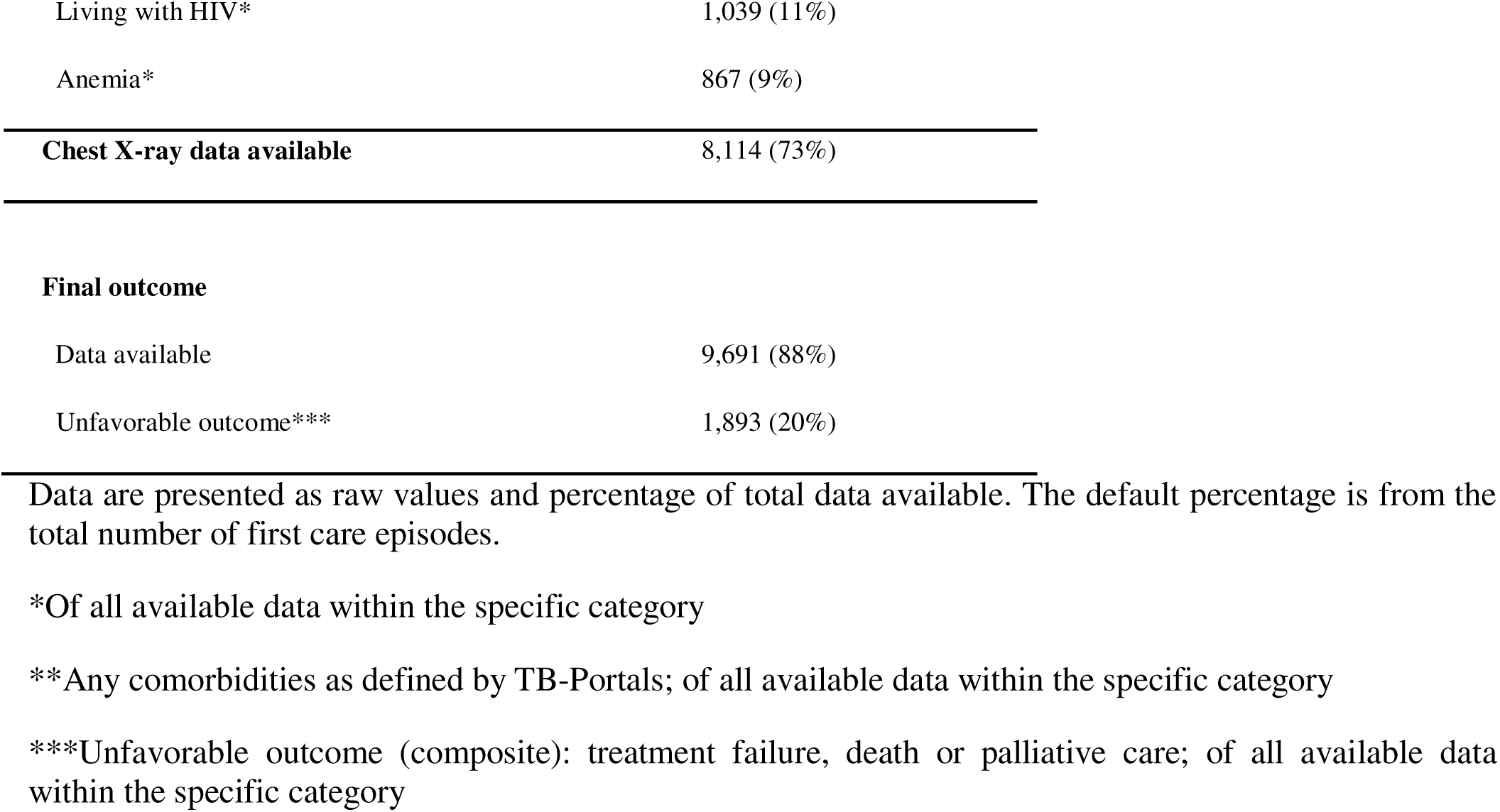
Description of accessed TB-Portals database.

**Supplementary Table 2.**
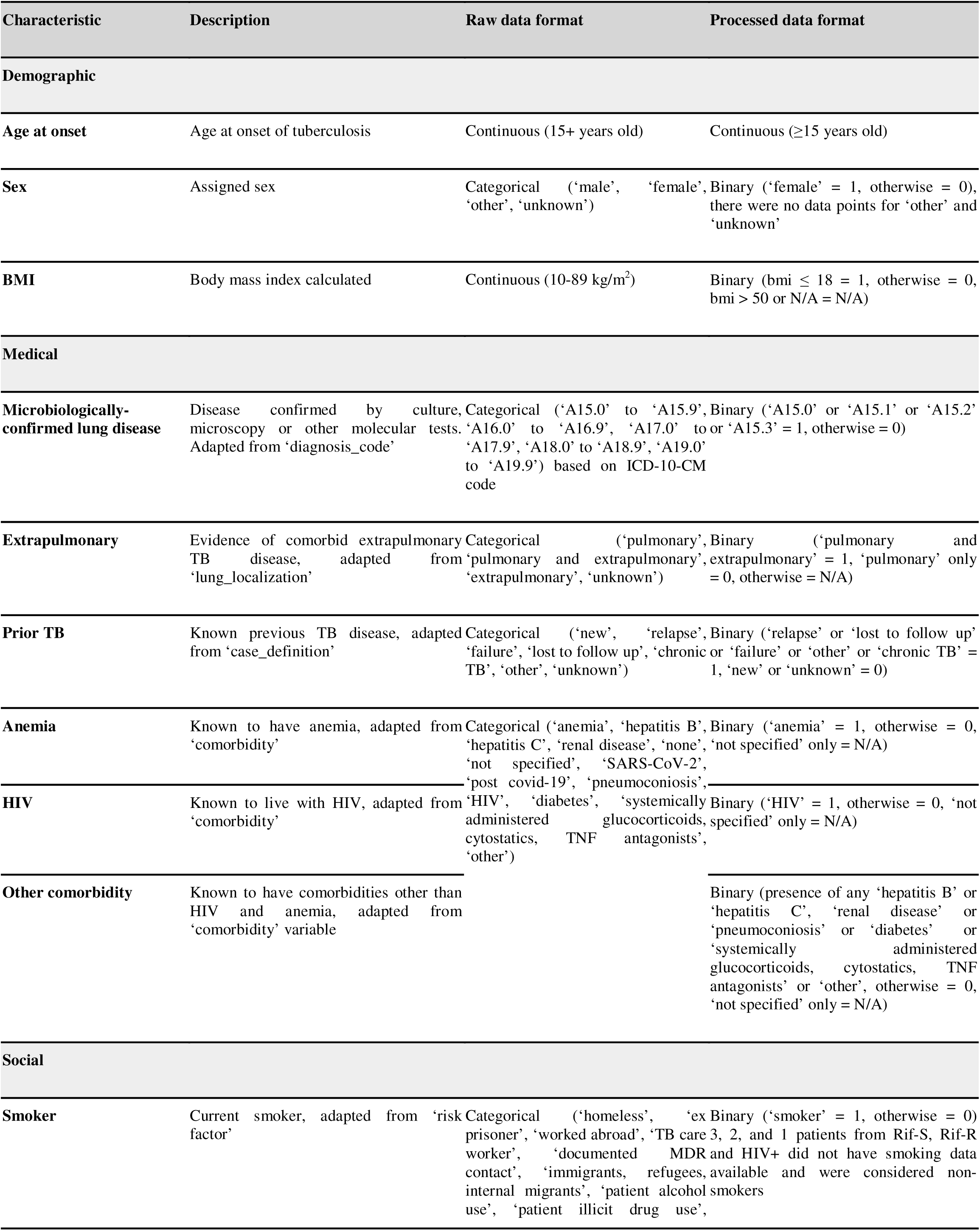
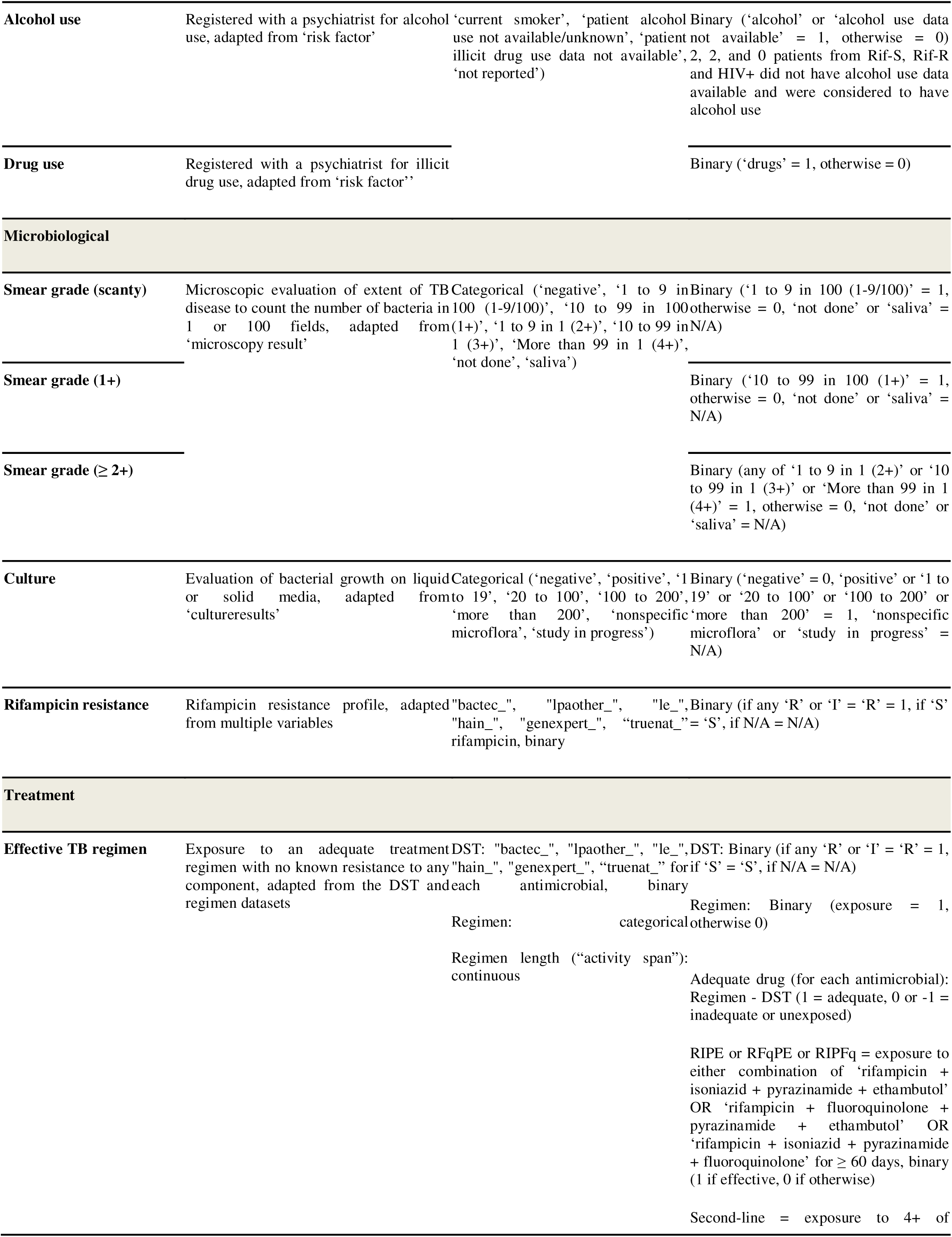
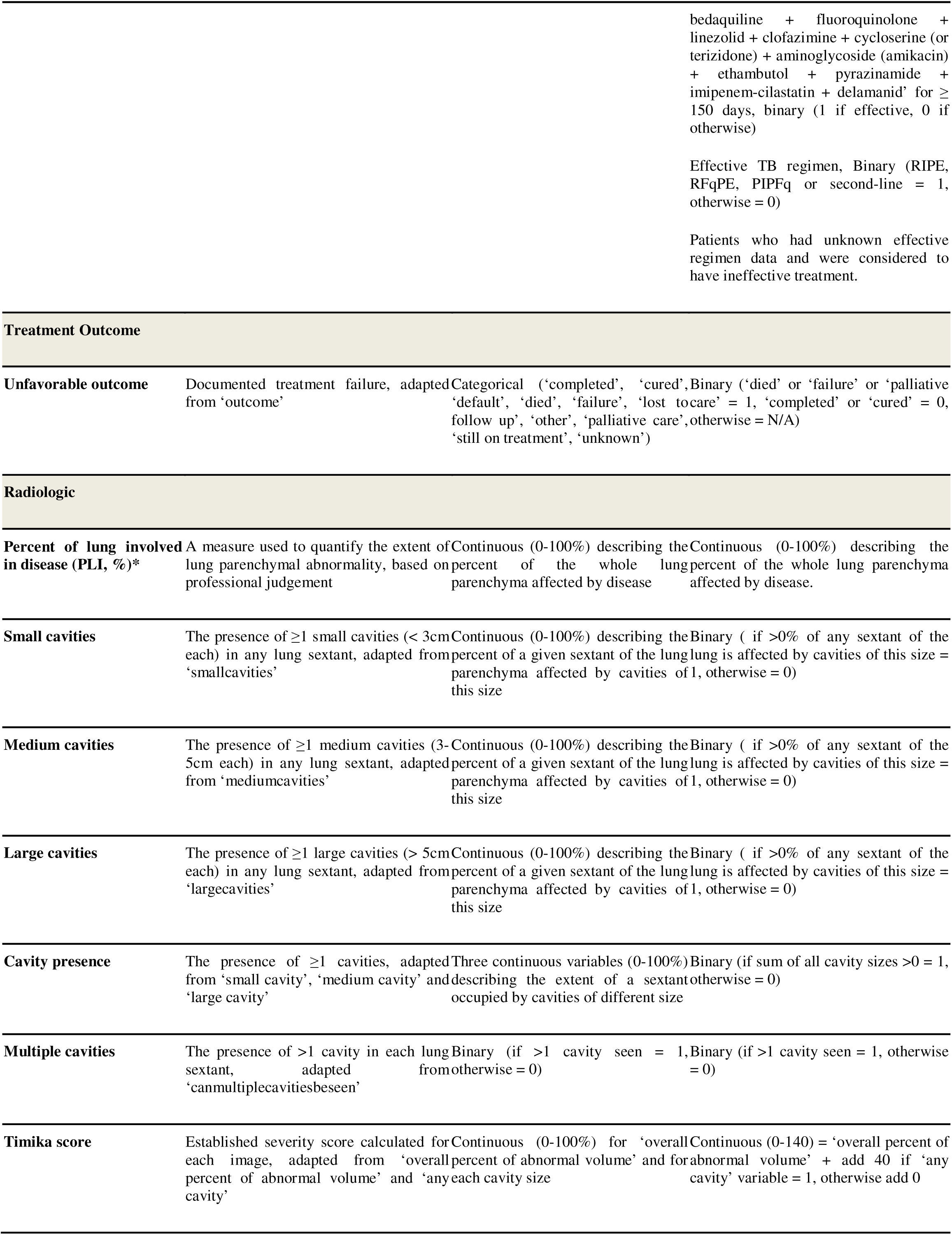
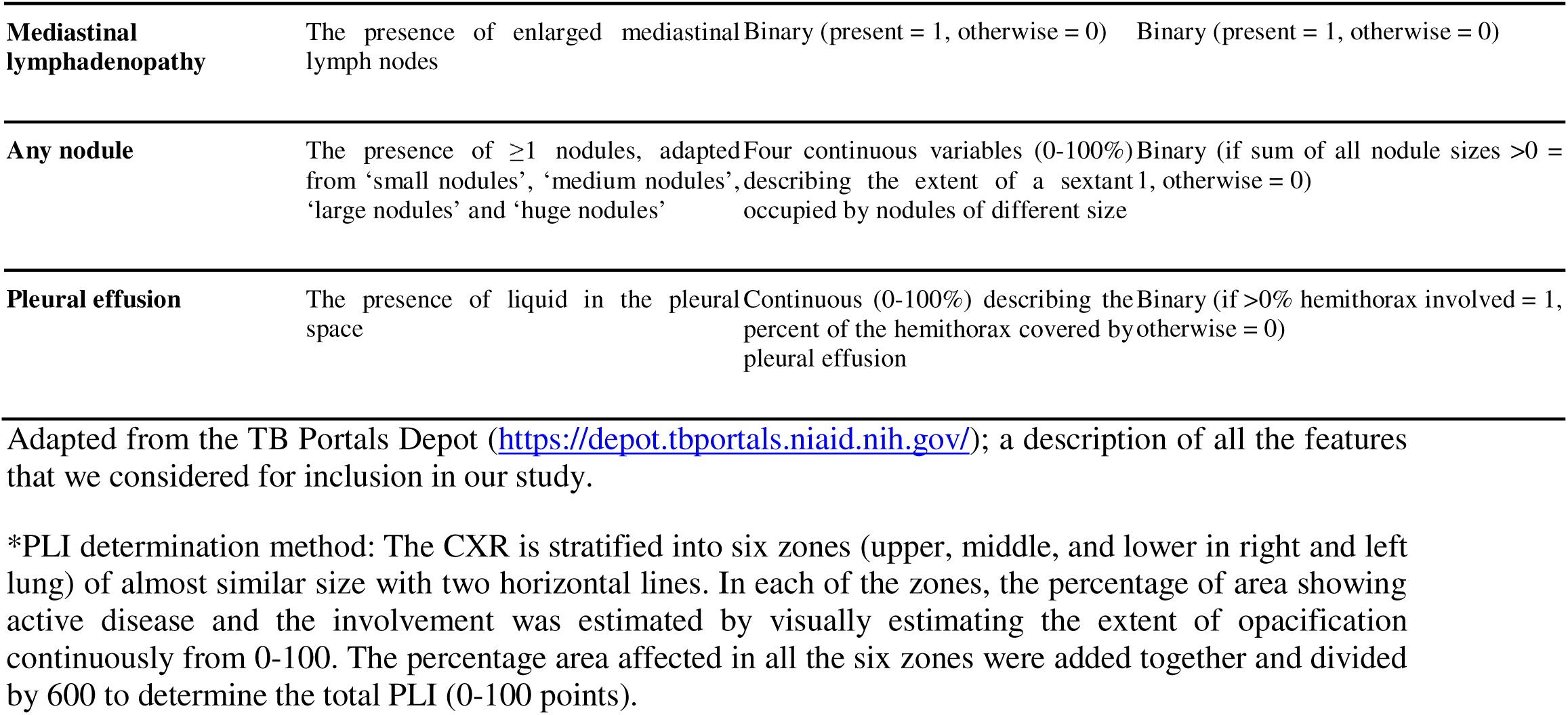
Data dictionary.

**Supplementary Table 3.**
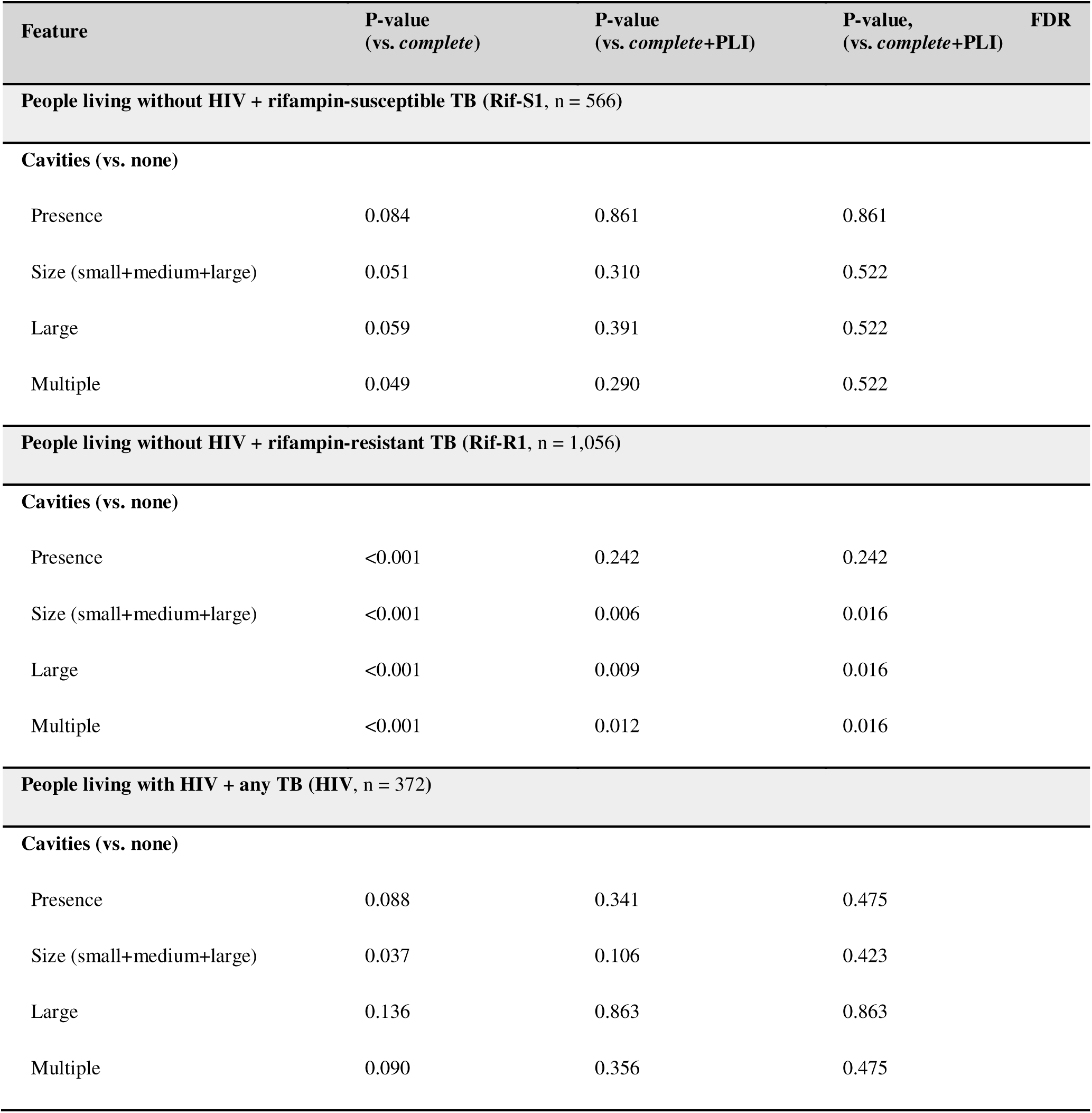
Addition of cavitary information to logistic regression models containing PLI. We built logistic regression models with and without PLI and the cavitary information of interest. We used likelihood ratio tests to assess significance of each feature added to the model based on the deviance method and generated p-values using the chi-squared method. We corrected for multiple hypothesis testing correction by adjusting the Benjamini-Hochberg false discovery rate at <0.05. *complete* = female (male as referent), age at onset of disease (continuous), BMI ≤ 18 kg/m^2^ (>18 kg/m^2^ as referent), extrapulmonary involvement, prior TB disease, anemia, other comorbidities (includes hepatic or renal disease, diabetes mellitus, immunosuppression, pneumoconiosis or other diseases), alcohol use, smoking, smear grade (scanty, 1+ or ≥ 2+ with negative microscopy as a referent), and effective drug therapy, *i.e.* RIPE or equivalent ≥ 60 days or second-line 4+ antimicrobials ≥ 150 days without known resistance to any of the components. Rifampicin resistance and antiretroviral use are added to the *complete* model for people living with HIV.

**Supplementary table 4.**
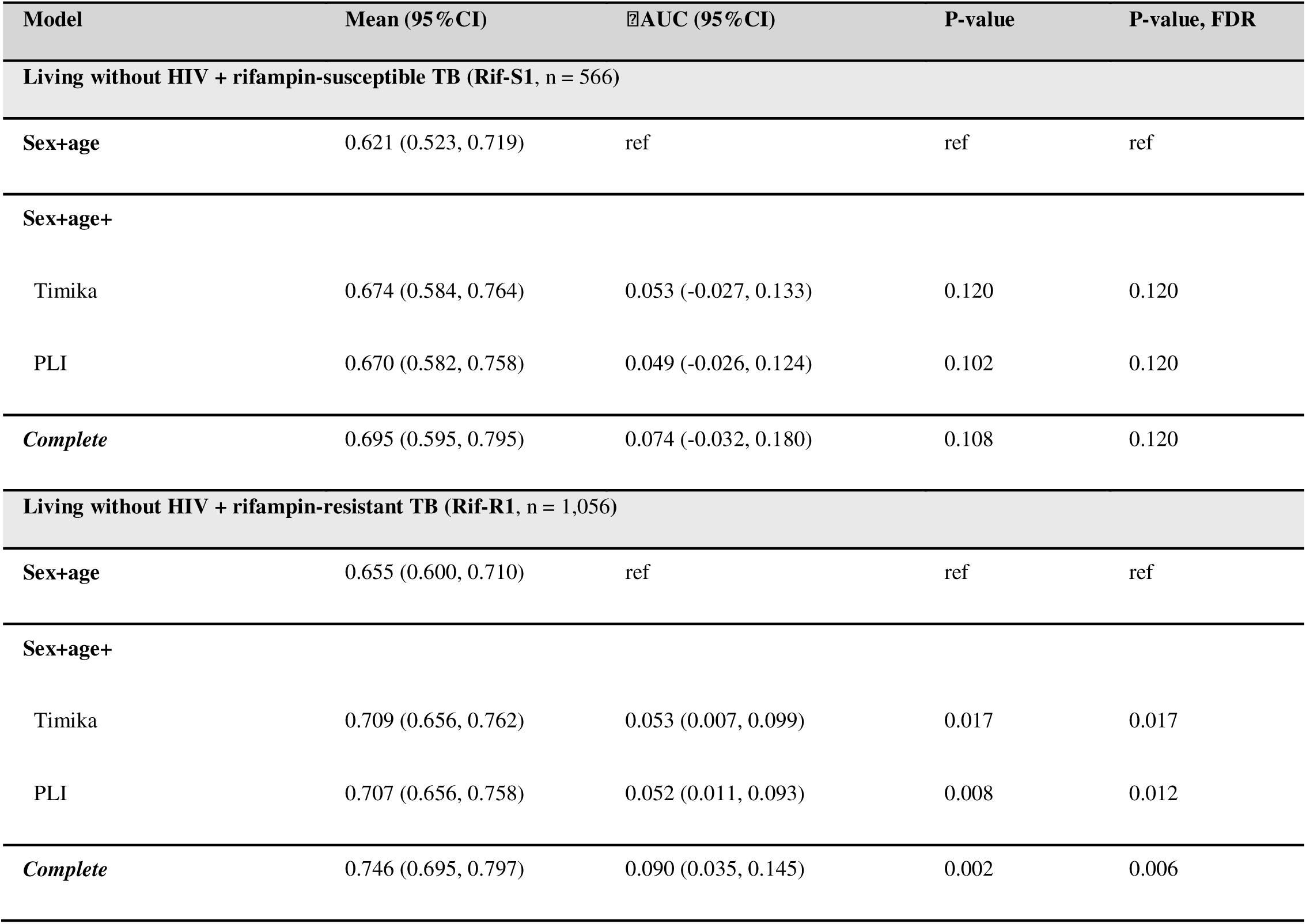
Subgroup analysis for predicting model accuracy based on rifampicin. We estimated the prediction accuracy using Rif-S1 (n = 566) or Rif-R1 (n = 1,056) for models built with (A) individual radiological features, (B) PLI + cavitary information and (C) non-radiological + radiological features. We used a Monte Carlo cross-validation approach with 1,000 iterations of resampling (75:25), trained on the 75% and tested of the 25% of each iteration. At every iteration, we computed the difference between model AUCs (ΔAUC), and the number of observed differences that were ≤ 0 were divided by the total number of observations to assess statistical significance using a one-tailed empirical p-value approach [p-value = (#ΔAUC) ≤0/1,000]. We corrected for multiple hypothesis testing by controlling the Benjamini-Hochberg false discovery rate to <0.05. IIAUC, Pval and FDR Pval were done compared to the sex+age model.

**Supplementary Table 5.**
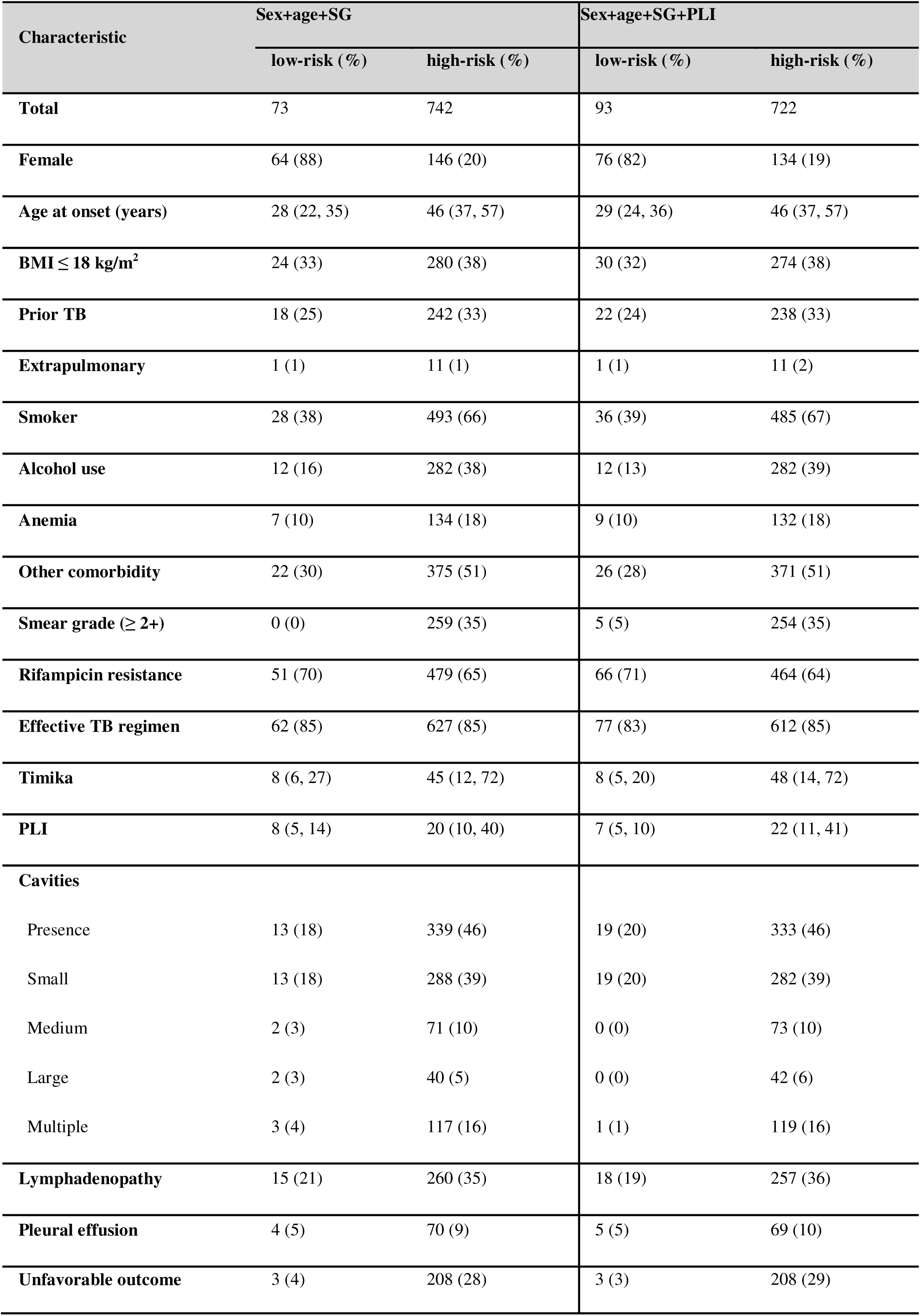
Low- and high-risk group baseline characteristics for the test data when separated based on the optimal threshold for each logistic regression model.

**Supplementary Table 6.**
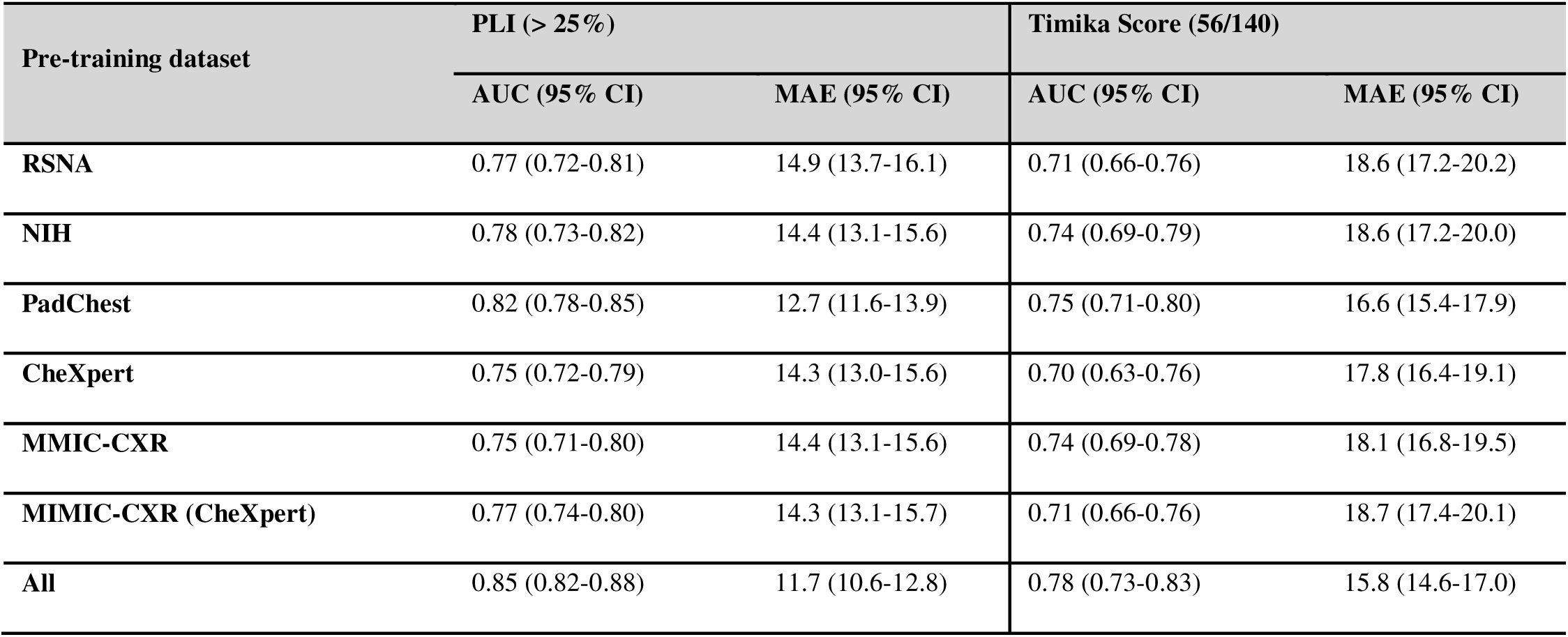
Machine learning model performance for prediction of PLI and Timika. The classifiers’ performance was assessed using the Area Under the Receiver Operating Characteristic Curve (AUC-ROC), accuracy, precision-recall curve, F1 score, sensitivity, and specificity. Bootstrapping was used to estimate the AUC by resampling predictions and labels from the test set 100 times, generating a distribution of AUC scores with 95% confidence intervals. For performance evaluation in the Regression analysis, Mean Squared Error (MSE), Mean Absolute Error (MAE), and R-squared (R²) score were used. Bootstrapping was applied 500 times to generate a distribution of these metrics, and 95% confidence intervals were calculated. Unfreeze last layer: trainable parameters 1,025.

