## Supplementary for "Tuberculosis disease severity assessment using clinical variables and radiology enabled by artificial intelligence"

**Materials and Methods**

**Methods for ML-based prediction of radiological features**

**a. Cohort selection and image quality check:**

Rif-S + Rif-R CXR

n = 7,213

CXR without outliers

n = 5,261

MONO2

n = 5,240

MONO1

n = 21

ML ready

MONO2

n = 5,261

**Supplementary Figures**

**Supplementary Figure 1. Geographic distribution of TB-Portals data.**

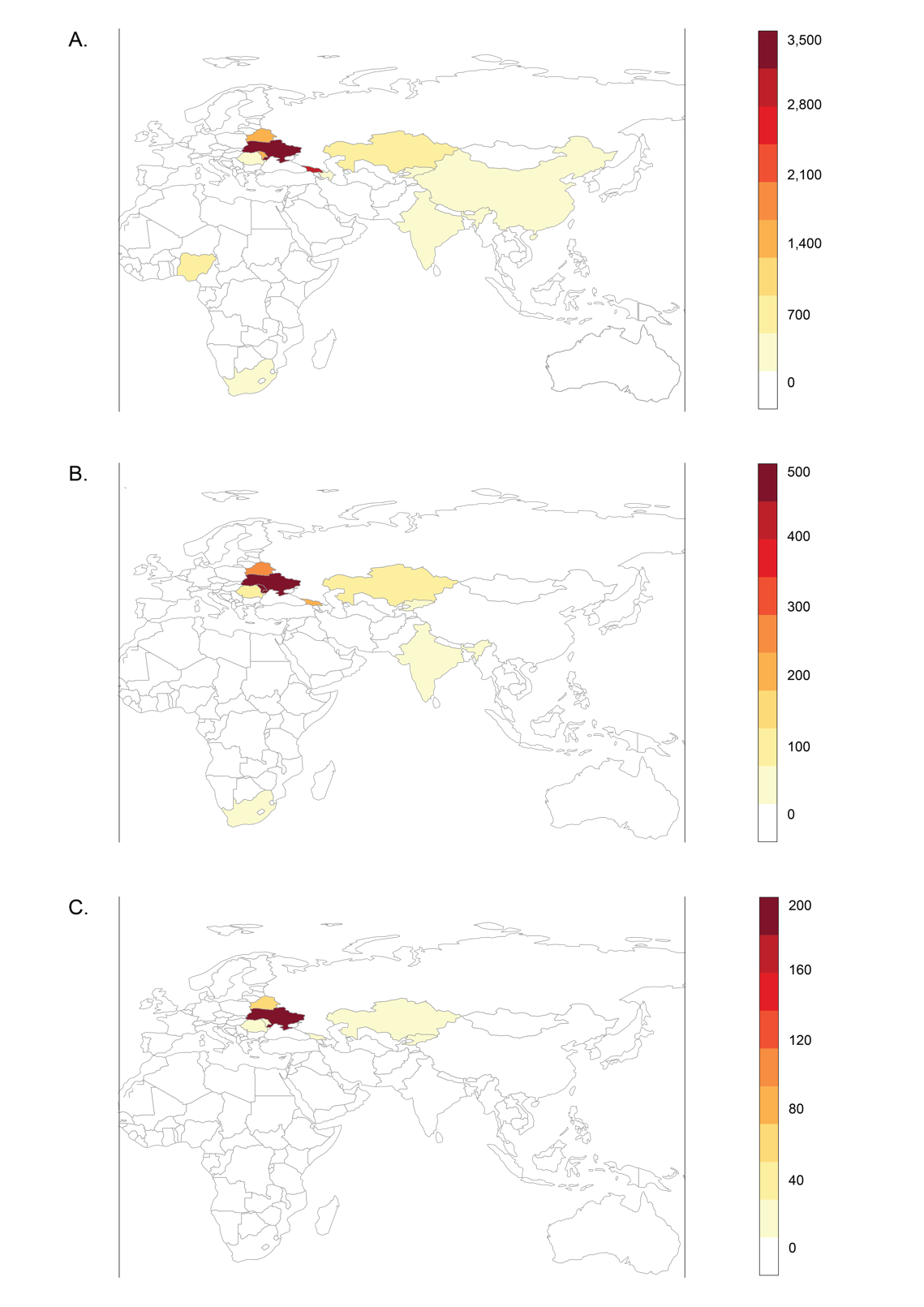

(A) Geographical distribution of all patients (first care episode) within the TB Portals database (n=11,067) with a scale ranging from 0-3,500 individuals. The dataset included patient cases from 13 countries: Ukraine, Georgia, Moldova, Belarus, Kazakhstan, Nigeria, Romania, Azerbaijan, South Africa, China, Kyrgyzstan, India, and Senegal. (B) Geographical distribution of cases included in the training-validation and HIV datasets (n=1,994) with a scale ranging from 0-500. (C) Geographical distribution of cases included in the test dataset (n=815) with a scale ranging from 0-200 individuals, respectively.

**Supplementary Figure 2. Examples of monochrome chest x-ray images present in the dataset.**

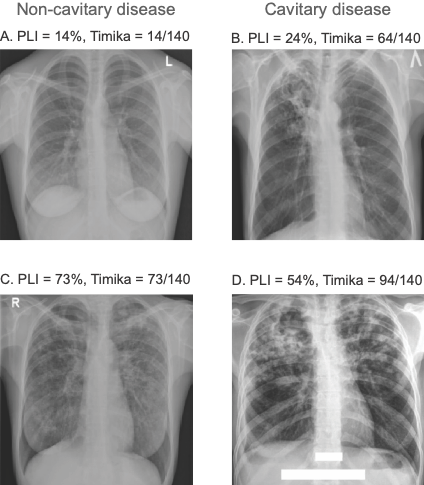

(A) low percent of lung involved in disease (PLI) and no cavitation, Timika score = PLI, (B) low PLI and cavitation, Timika score = PLI+40, (C) high PLI and no cavitation (Timika score = PLI) and (D) high PLI and cavitation, Timika score = PLI+40.

**Supplementary Figure 3. Prediction of unfavorable outcome using training data.**

A.    Radiological features alone

| **Model** | **Mean (95%CI)** | **𝝙AUC (95%CI)** | **P-value, FDR** |
| --- | --- | --- | --- |
| **PLI** | 0.656 (0.595, 0.717) | ref | ref |
| **Timika** | 0.655 (0.594, 0.716) | 0.001 (-0.029, 0.031) | 0.468 |
| **Cavities (vs. none)** |  |  |  |
| Presence | 0.583 (0.528, 0.638) | 0.073 (0.008, 0.138) | 0.027 |
| Size (small+medium+large) | 0.591 (0.532, 0.650) | 0.065 (0.000, 0.130) | 0.034 |
| Large | 0.547 (0.508, 0.586) | 0.109 (0.049, 0.169) | <0.001 |
| Multiple | 0.566 (0.519, 0.613) | 0.090 (0.030, 0.150) | <0.001 |
| **Lymphadenopathy** | 0.493 (0.448, 0.538) | 0.163 (0.089, 0.237) | <0.001 |

B.    Percent lung involved + cavities

| **Model** | **Mean (95%CI)** | **𝝙AUC (95%CI)** | **P-value, FDR** |
| --- | --- | --- | --- |
| **PLI** | 0.654 (0.593, 0.715) | ref | ref |
| **Timika** | 0.653 (0.590, 0.716) | 0.001 (-0.028, 0.030) | 0.586 |
| **PLI + Cavities** |  |  |  |
| Presence | 0.655 (0.592, 0.718) | -0.001 (-0.023, 0.021) | 0.586 |
| Size (small+medium+large) | 0.650 (0.587, 0.713) | 0.005 (-0.018, 0.028) | 0.586 |
| Large | 0.654 (0.593, 0.715) | -0.000 (-0.009, 0.009) | 0.586 |
| Multiple | 0.653 (0.592, 0.714) | 0.001 (-0.012, 0.014) | 0.586 |

C.    Non-radiological + radiological (training)

| **Model** | **Mean (95%CI)** | **𝝙AUC (95%CI)** | **P-value, FDR** |
| --- | --- | --- | --- |
| **Sex+age** | 0.655 (0.600, 0.710) | ref | ref |
| **Sex+age+** |  |  |  |
| Timika | 0.709 (0.656, 0.762) | 0.053 (0.007, 0.099) | 0.017 |
| PLI | 0.707 (0.656, 0.758) | 0.052 (0.011, 0.093) | 0.012 |
| ***Complete*** | 0.746 (0.695, 0.797) | 0.090 (0.035, 0.145) | 0.006 |

D.    KDE plots for model prediction accuracies

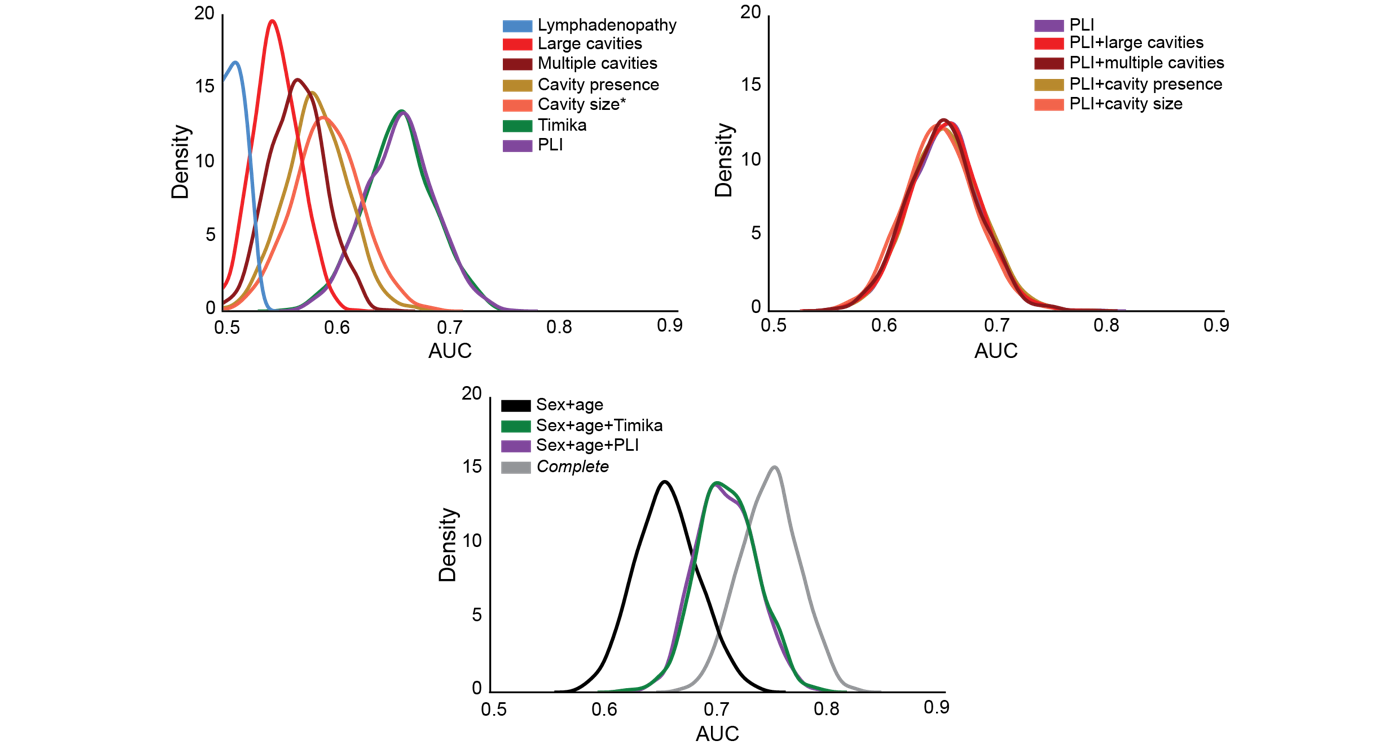

We estimated the prediction accuracy using Rif-S1 + Rif-R1 (n = 1,622) for models built with (A) individual radiological features, (B) PLI + cavitary information and (C) non-radiological + radiological features. We used a Monte Carlo cross-validation approach with 1,000 iterations of resampling (75:25), trained on the 75% (n = 1,216) and tested of the 25% (n = 406) of each iteration. At every iteration, we computed the difference between model AUCs (ΔAUC), and the number of observed differences that were ≤ 0 were divided by the total number of observations to assess statistical significance using a one-tailed empirical p-value approach [p-value = (#ΔAUC) ≤0/1,000]. We corrected for multiple hypothesis testing by controlling the Benjamini-Hochberg false discovery rate to <0.05. In (A) and (B), 𝝙AUC and FDR Pval were done compared to the PLI model, and in (C) 𝝙AUC and FDR Pval were done compared to the sex+age model. (D) The KDE plots are visual representations of the mean AUC and 95% confidence interval for individual radiological features (top left), PLI +/- cavitary information (top right) and the reduced model +/- PLI or Timika (bottom).

**Supplementary Figure 4. Prediction of unfavorable outcomes for radiological features with smear grade.**

A.

|  | **training-validation data** | | **Test data** | |
| --- | --- | --- | --- | --- |
| **Model** | **Mean (95%CI)** | **P-value, FDR*** | **Mean (95%CI)** | **P-value, FDR*** |
| **Sex+age+SG** | 0.683 (0.630, 0.736) | ref | 0.661 (0.622, 0.700) | ref |
| **Sex+age+SG+** |  |  |  |  |
| Timika | 0.713 (0.660, 0.766) | 0.057 | 0.673 (0.634, 0.712) | 0.188 |
| PLI | 0.714 (0.661, 0.767) | 0.057 | 0.689 (0.650, 0.728) | 0.004 |

B.

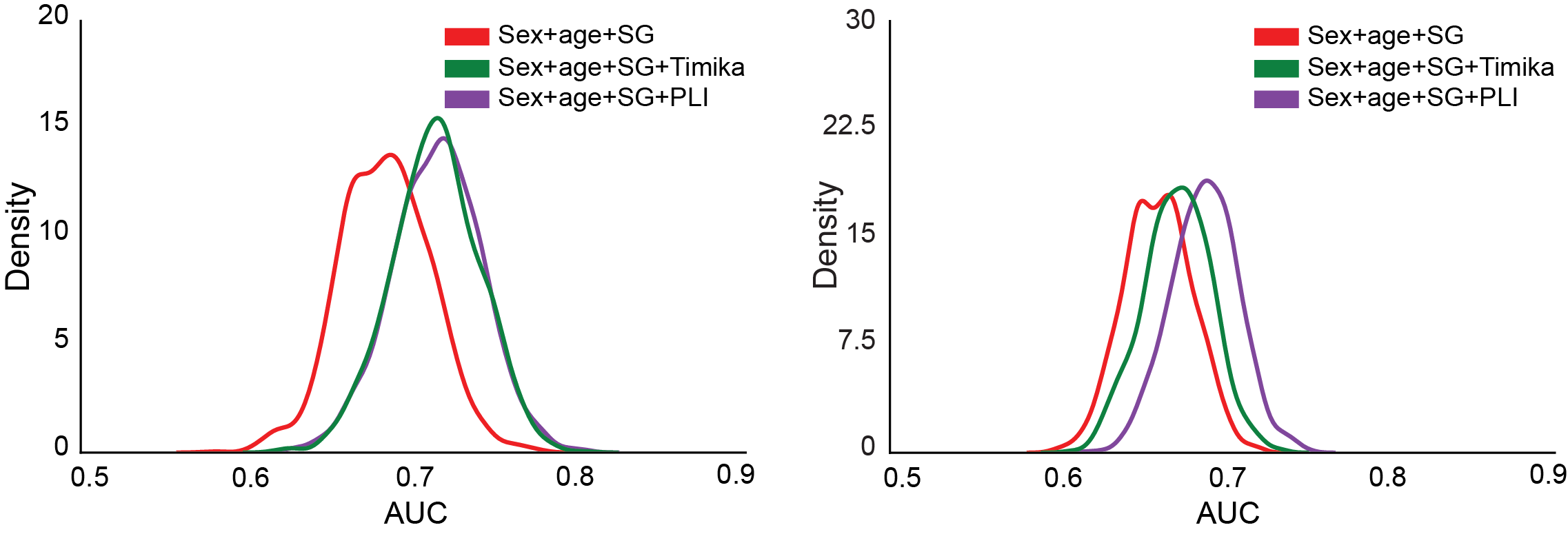

(A) We estimated the prediction accuracy for models built with sex+age+smear grade +/- PLI or Timika. For the training-validation prediction accuracy assessment, we used a Monte Carlo cross-validation approach with 1000 iterations of resampling (75:25), trained on the 75% (n = 1,216) and validated of the 25% (n = 406) of each iteration. For the test dataset, we trained the logistic regression models on Rif-S1 + Rif-R1 (n = 1,622) and predicted outcomes on Rif-S2 + Rif-R2 (n = 815). We used sampling with replacement (1,000 iterations) to generate a mean AUC and confidence intervals. The data represents the mean AUC of the 1000 iterations and the 95% CI (mean +/- 1.96 x standard deviation). At every iteration, we computed the difference between model AUCs (ΔAUC), and the number of observed differences that were ≤ 0 were divided by the total number of observations to assess statistical significance using a one-tailed empirical p-value approach [p-value = (#ΔAUC) ≤0/1,000]. We corrected for multiple hypothesis testing by controlling the Benjamini-Hochberg false discovery rate to <0.05. 𝝙AUC and FDR Pval were done compared to the sex+age+smear grade model.

(B) The KDE plots are visual representations of the mean AUC and 95% confidence interval for the asex+age+smear grade model +/- PLI or Timika for the training-validation analysis (left) and the test analysis (right).

**Supplementary Figure 5. Prediction of unfavorable outcomes for radiological characteristics in people living with HIV.**

A.

| **Model** | **Mean (95%CI)** | **𝝙AUC (95%CI)** | **P-value, FDR** |
| --- | --- | --- | --- |
| **Sex+age** | 0.522 (0.432, 0.612) | ref | ref |
| **Timika** | 0.600 (0.529, 0.671) | 0.078 (-0.038, 0.194) | 0.080 |
| **PLI** | 0.596 (0.522, 0.670) | 0.074 (-0.043, 0.191) | 0.080 |
| **Sex+age+** |  |  |  |
| Timika | 0.581 (0.508, 0.654) | 0.059 (-0.027, 0.145) | 0.080 |
| PLI | 0.572 (0.498, 0.646) | 0.050 (-0.029, 0.129) | 0.080 |
| ***Complete*** | 0.704 (0.637, 0.771) | 0.182 (0.084, 0.280) | <0.001 |

B.

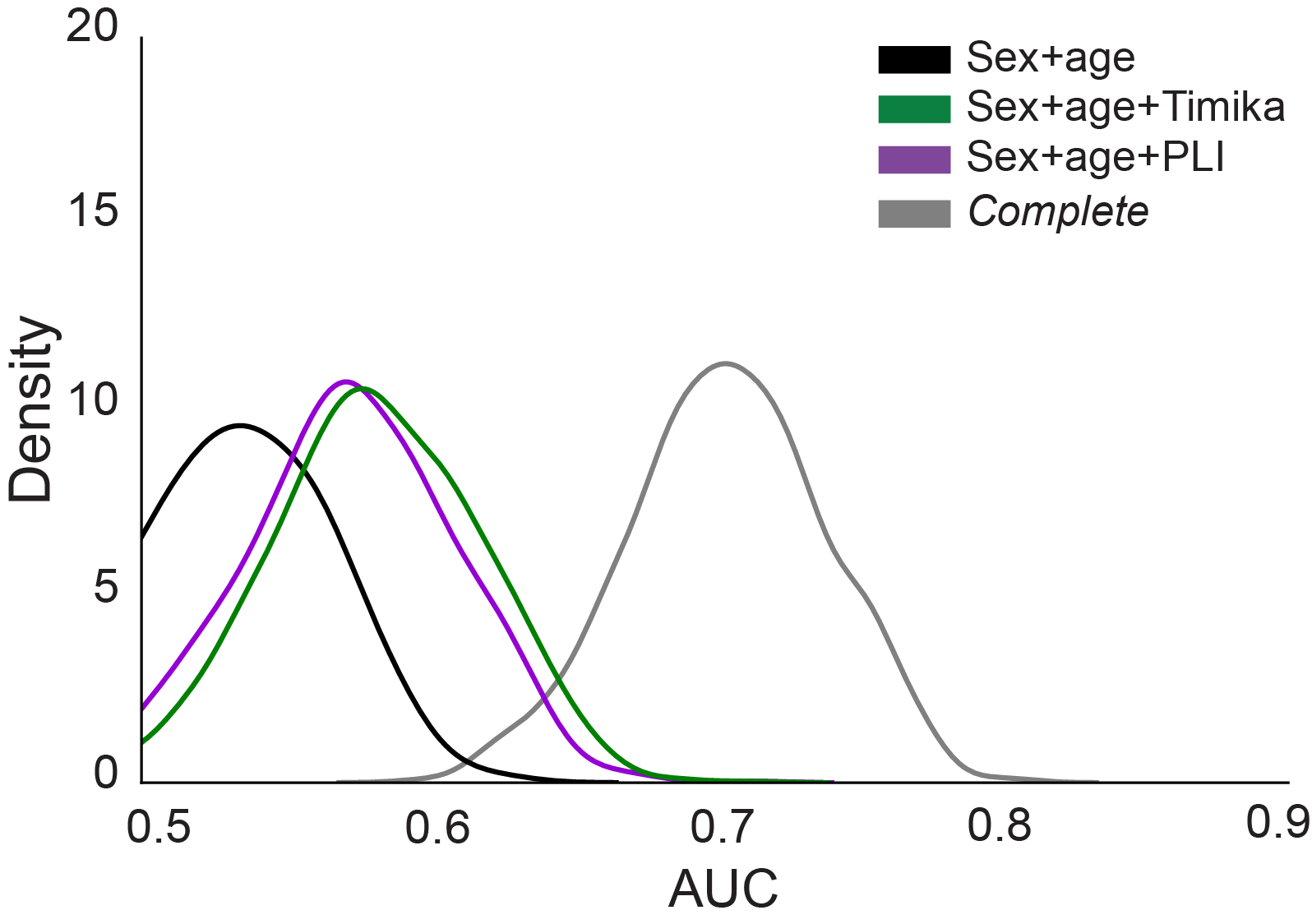

We estimated the prediction accuracy using HIV (n = 372). We used a Monte Carlo cross-validation approach with 1000 iterations of resampling (75:25), trained on the 75% (n = 223) and validated of the 25% (n = 149) of each iteration.

(A) At every iteration, we computed the difference between model AUCs (ΔAUC). The data represents the mean AUC of the 1000 iterations and the 95% CI. The number of observed differences that were ≤ 0 were divided by the total number of observations to assess statistical significance using a one-tailed empirical p-value approach [P-value = (#ΔAUC) ≤0/1,000]. We corrected for multiple hypothesis testing by controlling the Benjamini-Hochberg false discovery rate to <0.05. 𝝙AUC and the FDR-corrected P-value were done compared to the sex+age model.

(B) The KDE plots are visual representations of the mean AUC and 95% confidence interval for the reduced model +/- PLI or Timika.

**Supplementary Figure 6. Testing SPECTRA with modifications for measuring severity in Rif-S and endTB-Q with modifications for measuring severity in Rif-R.**

A.

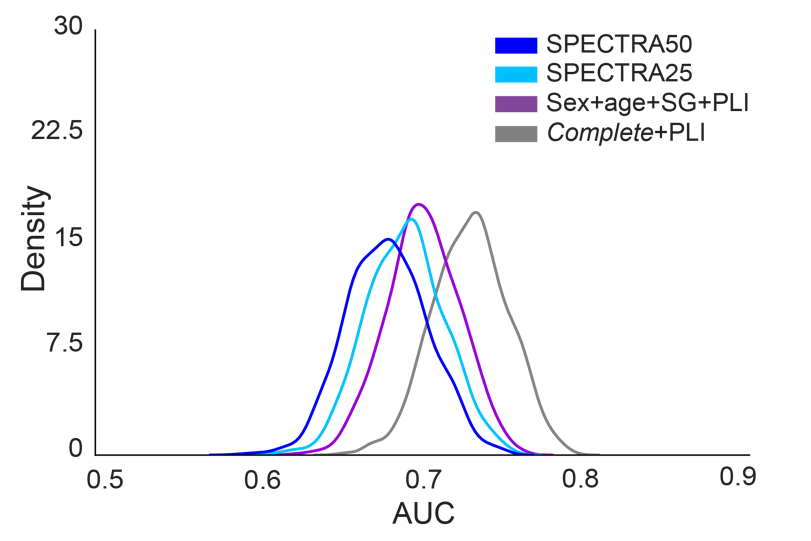

B.

| **Model** | **Mean (95%CI)** | **Mean 𝝙AUC (95%CI)** | **FDR Pval** |
| --- | --- | --- | --- |
| **SPECTRA50** | 0.678 (0.629, 0.727) | ref | ref |
| **SPECTRA25** | 0.689 (0.642, 0.736) | 0.011 (-0.010, 0.032) | 0.164 |
| **Sex+age+SG+PLI** | 0.700 (0.657, 0.743) | 0.022 (-0.022, 0.066) | 0.164 |
| ***Complete*+PLI** | 0.729 (0.684, 0.774) | 0.051 (0.011, 0.091) | 0.015 |

C.	D.

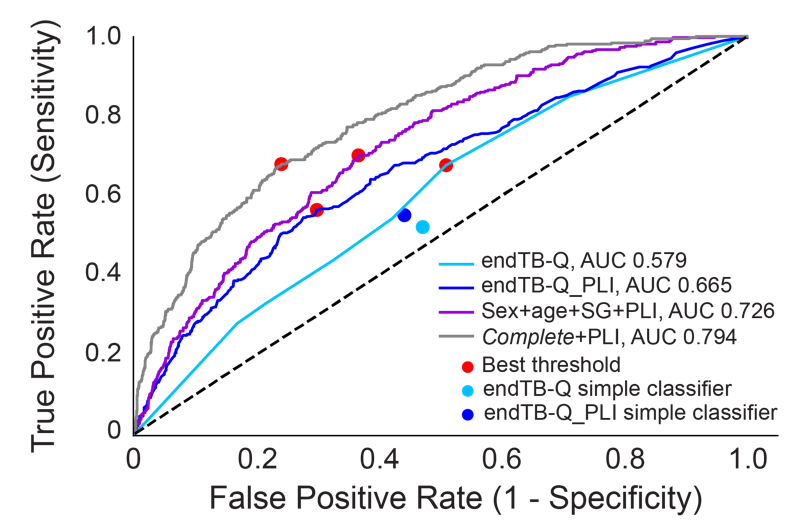

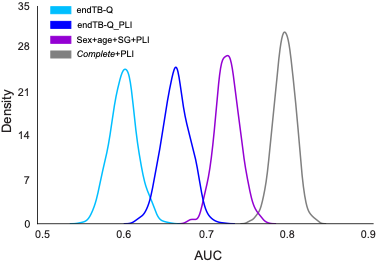

E.

| **Model** | **Mean (95%CI)** | **Mean 𝝙AUC (95%CI)** | **FDR Pval** |
| --- | --- | --- | --- |
| **endTB-Q** | 0.602 (0.571, 0.633) | ref | ref |
| **endTB-Q_PLI** | 0.665 (0.634, 0.696) | 0.063 (0.035, 0.091) | <0.001 |
| **Sex+age+SG+PLI** | 0.726 (0.699, 0.753) | 0.124 (0.091, 0.157) | <0.001 |
| ***Complete*+PLI** | 0.794 (0.769, 0.819) | 0.192 (0.159, 0.225) | <0.001 |

**Supplementary Tables**

**Supplementary Table 1. Description of accessed TB-Portals database.**

| **Characteristic** | **Frequency** |
| --- | --- |
| **Cases** |  |
| All care episodes | 11,282 |
| First care episode | 11,067 (100%) |
| **Country** |  |
| Ukraine | 3,176 (29%) |
| Georgia | 2,953 (27%) |
| Moldova | 1,280 (12%) |
| Other | 3,658 (33%) |
| **Female** | 2,994 (27%) |
| **Age at onset (median and IQR)** | 41 (32, 52) |
| **BMI** |  |
| Data available | 7,885 (71%) |
| Median and IQR* | 20.3 (18.2, 22.5) |
| **Prior TB** | 3,706 (33%) |
| **Pulmonary TB** | 10,802 (98%) |
| **Resistance profile** |  |
| Pan-susceptible | 3,719 (34%) |
| Mono- or poly-resistant | 1,092 (10%) |
| Multidrug-resistant | 6,255 (57%) |
| **Comorbidity** |  |
| Data available | 9,407 (85%) |
| Any comorbidity** | 4,468 (47%) |
| Living with HIV* | 1,039 (11%) |
| Anemia* | 867 (9%) |
| **Chest X-ray data available** | 8,114 (73%) |
| **Final outcome** |  |
| Data available | 9,691 (88%) |
| Unfavorable outcome*** | 1,893 (20%) |

Data are presented as raw values and percentage of total data available. The default percentage is from the total number of first care episodes.

*Of all available data within the specific category

**Any comorbidities as defined by TB-Portals; of all available data within the specific category

***Unfavorable outcome (composite): treatment failure, death or palliative care; of all available data within the specific category

**Supplementary Table 2. Data dictionary.**

| **Characteristic** | **Description** | **Raw data format** | **Processed data format** |
| --- | --- | --- | --- |
| **Demographic** | | | |
| **Age at onset** | Age at onset of tuberculosis | Continuous (15+ years old) | Continuous (≥15 years old) |
| **Sex** | Assigned sex | Categorical (‘male’, ‘female’, ‘other’, ‘unknown’) | Binary (‘female’ = 1, otherwise = 0), there were no data points for ‘other’ and ‘unknown’ |
| **BMI** | Body mass index calculated | Continuous (10-89 kg/m^2^) | Binary (bmi ≤ 18 = 1, otherwise = 0, bmi > 50 or N/A = N/A) |
| **Medical** | | | |
| **Microbiologically-confirmed lung disease** | Disease confirmed by culture, microscopy or other molecular tests. Adapted from ‘diagnosis_code’ | Categorical (‘A15.0’ to ‘A15.9’, ‘A16.0’ to ‘A16.9’, ‘A17.0’ to ‘A17.9’, ‘A18.0’ to ‘A18.9’, ‘A19.0’ to ‘A19.9’) based on ICD-10-CM code | Binary (‘A15.0’ or ‘A15.1’ or ‘A15.2’ or ‘A15.3’ = 1, otherwise = 0) |
| **Extrapulmonary** | Evidence of comorbid extrapulmonary TB disease, adapted from ‘lung_localization’ | Categorical (‘pulmonary’, ‘pulmonary and extrapulmonary’, ‘extrapulmonary’, ‘unknown’) | Binary (‘pulmonary and extrapulmonary’ = 1, ‘pulmonary’ only = 0, otherwise = N/A) |
| **Prior TB** | Known previous TB disease, adapted from ‘case_definition’ | Categorical (‘new’, ‘relapse’, ‘failure’, ‘lost to follow up’, ‘chronic TB’, ‘other’, ‘unknown’) | Binary (‘relapse’ or ‘lost to follow up’ or ‘failure’ or ‘other’ or ‘chronic TB’ = 1, ‘new’ or ‘unknown’ = 0) |
| **Anemia** | Known to have anemia, adapted from ‘comorbidity’ | Categorical (‘anemia’, ‘hepatitis B’, ‘hepatitis C’, ‘renal disease’, ‘none’, ‘not specified’, ‘SARS-CoV-2’, ‘post covid-19’, ‘pneumoconiosis’, ‘HIV’, ‘diabetes’, ‘systemically administered glucocorticoids, cytostatics, TNF antagonists’, ‘other’) | Binary (‘anemia’ = 1, otherwise = 0, ‘not specified’ only = N/A) |
| **HIV** | Known to live with HIV, adapted from ‘comorbidity’ |  | Binary (‘HIV’ = 1, otherwise = 0, ‘not specified’ only = N/A) |
| **Other comorbidity** | Known to have comorbidities other than HIV and anemia, adapted from ‘comorbidity’ variable |  | Binary (presence of any ‘hepatitis B’ or ‘hepatitis C’, ‘renal disease’ or ‘pneumoconiosis’ or ‘diabetes’ or ‘systemically administered glucocorticoids, cytostatics, TNF antagonists’ or ‘other’, otherwise = 0, ‘not specified’ only = N/A) |
| **Social** | | | |
| **Smoker** | Current smoker, adapted from ‘risk factor’ | Categorical (‘homeless’, ‘ex prisoner’, ‘worked abroad’, ‘TB care worker’, ‘documented MDR contact’, ‘immigrants, refugees, internal migrants’, ‘patient alcohol use’, ‘patient illicit drug use’, ‘current smoker’, ‘patient alcohol use not available/unknown’, ‘patient illicit drug use data not available’, ‘not reported’) | Binary (‘smoker’ = 1, otherwise = 0) 3, 2, and 1 patients from Rif-S, Rif-R and HIV+ did not have smoking data available and were considered non-smokers |
| **Alcohol use** | Registered with a psychiatrist for alcohol use, adapted from ‘risk factor’ |  | Binary (‘alcohol’ or ‘alcohol use data not available’ = 1, otherwise = 0) 2, 2, and 0 patients from Rif-S, Rif-R and HIV+ did not have alcohol use data available and were considered to have alcohol use |
| **Drug use** | Registered with a psychiatrist for illicit drug use, adapted from ‘risk factor’’ |  | Binary (‘drugs’ = 1, otherwise = 0) |
| **Microbiological** | | | |
| **Smear grade (scanty)** | Microscopic evaluation of extent of TB disease to count the number of bacteria in 1 or 100 fields, adapted from ‘microscopy result’ | Categorical (‘negative’, ‘1 to 9 in 100 (1-9/100)’, ‘10 to 99 in 100 (1+)’, ‘1 to 9 in 1 (2+)’, ‘10 to 99 in 1 (3+)’, ‘More than 99 in 1 (4+)’, ‘not done’, ‘saliva’) | Binary (‘1 to 9 in 100 (1-9/100)’ = 1, otherwise = 0, ‘not done’ or ‘saliva’ = N/A) |
| **Smear grade (1+)** |  |  | Binary (‘10 to 99 in 100 (1+)’ = 1, otherwise = 0, ‘not done’ or ‘saliva’ = N/A) |
| **Smear grade (≥ 2+)** |  |  | Binary (any of ‘1 to 9 in 1 (2+)’ or ‘10 to 99 in 1 (3+)’ or ‘More than 99 in 1 (4+)’ = 1, otherwise = 0, ‘not done’ or ‘saliva’ = N/A) |
| **Culture** | Evaluation of bacterial growth on liquid or solid media, adapted from ‘cultureresults’ | Categorical (‘negative’, ‘positive’, ‘1 to 19’, ‘20 to 100’, ‘100 to 200’, ‘more than 200’, ‘nonspecific microflora’, ‘study in progress’) | Binary (‘negative’ = 0, ‘positive’ or ‘1 to 19’ or ‘20 to 100’ or ‘100 to 200’ or ‘more than 200’ = 1, ‘nonspecific microflora’ or ‘study in progress’ = N/A) |
| **Rifampicin resistance** | Rifampicin resistance profile, adapted from multiple variables | "bactec_", "lpaother_", "le_", "hain_", "genexpert_", “truenat_” rifampicin, binary | Binary (if any ‘R’ or ‘I’ = ‘R’ = 1, if ‘S’ = ‘S’, if N/A = N/A) |
| **Treatment** | | | |
| **Effective TB regimen** | Exposure to an adequate treatment regimen with no known resistance to any component, adapted from the DST and regimen datasets | DST: "bactec_", "lpaother_", "le_", "hain_", "genexpert_", “truenat_” for each antimicrobial, binary  Regimen: categorical  Regimen length (“activity span”): continuous | DST: Binary (if any ‘R’ or ‘I’ = ‘R’ = 1, if ‘S’ = ‘S’, if N/A = N/A)  Regimen: Binary (exposure = 1, otherwise 0)  Adequate drug (for each antimicrobial): Regimen - DST (1 = adequate, 0 or -1 = inadequate or unexposed)  RIPE or RFqPE or RIPFq = exposure to either combination of ‘rifampicin + isoniazid + pyrazinamide + ethambutol’ OR ‘rifampicin + fluoroquinolone + pyrazinamide + ethambutol’ OR ‘rifampicin + isoniazid + pyrazinamide + fluoroquinolone’ for ≥ 60 days, binary (1 if effective, 0 if otherwise)  Second-line = exposure to 4+ of bedaquiline + fluoroquinolone + linezolid + clofazimine + cycloserine (or terizidone) + aminoglycoside (amikacin) + ethambutol + pyrazinamide + imipenem-cilastatin + delamanid’ for ≥ 150 days, binary (1 if effective, 0 if otherwise)  Effective TB regimen, Binary (RIPE, RFqPE, PIPFq or second-line = 1, otherwise = 0)  Patients who had unknown effective regimen data and were considered to have ineffective treatment. |
| **Treatment Outcome** | | | |
| **Unfavorable outcome** | Documented treatment failure, adapted from ‘outcome’ | Categorical (‘completed’, ‘cured’, ‘default’, ‘died’, ‘failure’, ‘lost to follow up’, ‘other’, ‘palliative care’, ‘still on treatment’, ‘unknown’) | Binary (‘died’ or ‘failure’ or ‘palliative care’ = 1, ‘completed’ or ‘cured’ = 0, otherwise = N/A) |
| **Radiologic** | | | |
| **Percent of lung involved in disease (PLI, %)*** | A measure used to quantify the extent of lung parenchymal abnormality, based on professional judgement | Continuous (0-100%) describing the percent of the whole lung parenchyma affected by disease | Continuous (0-100%) describing the percent of the whole lung parenchyma affected by disease. |
| **Small cavities** | The presence of ≥1 small cavities (< 3cm each) in any lung sextant, adapted from ‘smallcavities’ | Continuous (0-100%) describing the percent of a given sextant of the lung parenchyma affected by cavities of this size | Binary ( if >0% of any sextant of the lung is affected by cavities of this size = 1, otherwise = 0) |
| **Medium cavities** | The presence of ≥1 medium cavities (3-5cm each) in any lung sextant, adapted from ‘mediumcavities’ | Continuous (0-100%) describing the percent of a given sextant of the lung parenchyma affected by cavities of this size | Binary ( if >0% of any sextant of the lung is affected by cavities of this size = 1, otherwise = 0) |
| **Large cavities** | The presence of ≥1 large cavities (> 5cm each) in any lung sextant, adapted from ‘largecavities’ | Continuous (0-100%) describing the percent of a given sextant of the lung parenchyma affected by cavities of this size | Binary ( if >0% of any sextant of the lung is affected by cavities of this size = 1, otherwise = 0) |
| **Cavity presence** | The presence of ≥1 cavities, adapted from ‘small cavity’, ‘medium cavity’ and ‘large cavity’ | Three continuous variables (0-100%) describing the extent of a sextant occupied by cavities of different size | Binary (if sum of all cavity sizes >0 = 1, otherwise = 0) |
| **Multiple cavities** | The presence of >1 cavity in each lung sextant, adapted from ‘canmultiplecavitiesbeseen’ | Binary (if >1 cavity seen = 1, otherwise = 0) | Binary (if >1 cavity seen = 1, otherwise = 0) |
| **Timika score** | Established severity score calculated for each image, adapted from ‘overall percent of abnormal volume’ and ‘any cavity’ | Continuous (0-100%) for ‘overall percent of abnormal volume’ and for each cavity size | Continuous (0-140) = ‘overall percent of abnormal volume’ + add 40 if ‘any cavity’ variable = 1, otherwise add 0 |
| **Mediastinal lymphadenopathy** | The presence of enlarged mediastinal lymph nodes | Binary (present = 1, otherwise = 0) | Binary (present = 1, otherwise = 0) |
| **Any nodule** | The presence of ≥1 nodules, adapted from ‘small nodules’, ‘medium nodules’, ‘large nodules’ and ‘huge nodules’ | Four continuous variables (0-100%) describing the extent of a sextant occupied by nodules of different size | Binary (if sum of all nodule sizes >0 = 1, otherwise = 0) |
| **Pleural effusion** | The presence of liquid in the pleural space | Continuous (0-100%) describing the percent of the hemithorax covered by pleural effusion | Binary (if >0% hemithorax involved = 1, otherwise = 0) |

Adapted from the TB Portals Depot (<https://depot.tbportals.niaid.nih.gov/>); a description of all the features that we considered for inclusion in our study.

*PLI determination method: The CXR is stratified into six zones (upper, middle, and lower in right and left lung) of almost similar size with two horizontal lines. In each of the zones, the percentage of area showing active disease and the involvement was estimated by visually estimating the extent of opacification continuously from 0-100. The percentage area affected in all the six zones were added together and divided by 600 to determine the total PLI (0-100 points).

**Supplementary Table 3. Addition of cavitary information to logistic regression models containing PLI.**

| **Feature** | **P-value  (vs. *complete*)** | **P-value  (vs. *complete*+PLI)** | **P-value, FDR  (vs. *complete*+PLI)** |
| --- | --- | --- | --- |
| **People living without HIV + rifampin-susceptible TB (Rif-S1**, n = 566**)** | | | |
| **Cavities (vs. none)** |  |  |  |
| Presence | 0.084 | 0.861 | 0.861 |
| Size (small+medium+large) | 0.051 | 0.310 | 0.522 |
| Large | 0.059 | 0.391 | 0.522 |
| Multiple | 0.049 | 0.290 | 0.522 |
| **People living without HIV + rifampin-resistant TB (Rif-R1**, n = 1,056**)** | | | |
| **Cavities (vs. none)** |  |  |  |
| Presence | <0.001 | 0.242 | 0.242 |
| Size (small+medium+large) | <0.001 | 0.006 | 0.016 |
| Large | <0.001 | 0.009 | 0.016 |
| Multiple | <0.001 | 0.012 | 0.016 |
| **People living with HIV + any TB (HIV**, n = 372**)** | | | |
| **Cavities (vs. none)** |  |  |  |
| Presence | 0.088 | 0.341 | 0.475 |
| Size (small+medium+large) | 0.037 | 0.106 | 0.423 |
| Large | 0.136 | 0.863 | 0.863 |
| Multiple | 0.090 | 0.356 | 0.475 |

We built logistic regression models with and without PLI and the cavitary information of interest. We used likelihood ratio tests to assess significance of each feature added to the model based on the deviance method and generated p-values using the chi-squared method. We corrected for multiple hypothesis testing correction by adjusting the Benjamini-Hochberg false discovery rate at <0.05. *complete* = female (male as referent), age at onset of disease (continuous), BMI ≤ 18 kg/m^2^ (>18 kg/m^2^ as referent), extrapulmonary involvement, prior TB disease, anemia, other comorbidities (includes hepatic or renal disease, diabetes mellitus, immunosuppression, pneumoconiosis or other diseases), alcohol use, smoking, smear grade (scanty, 1+ or ≥ 2+ with negative microscopy as a referent), and effective drug therapy, *i.e.* RIPE or equivalent ≥ 60 days or second-line 4+ antimicrobials ≥ 150 days without known resistance to any of the components. Rifampicin resistance and antiretroviral use are added to the *complete* model for people living with HIV.

**Supplementary table 4. Subgroup analysis for predicting model accuracy based on rifampicin resistance.**

| **Model** | **Mean (95%CI)** | **𝝙AUC (95%CI)** | **P-value** | **P-value, FDR** |
| --- | --- | --- | --- | --- |
| **Living without HIV + rifampin-susceptible TB (Rif-S1**, n = 566**)** | | | | |
| **Sex+age** | 0.621 (0.523, 0.719) | ref | ref | ref |
| **Sex+age+** |  |  |  |  |
| Timika | 0.674 (0.584, 0.764) | 0.053 (-0.027, 0.133) | 0.120 | 0.120 |
| PLI | 0.670 (0.582, 0.758) | 0.049 (-0.026, 0.124) | 0.102 | 0.120 |
| ***Complete*** | 0.695 (0.595, 0.795) | 0.074 (-0.032, 0.180) | 0.108 | 0.120 |
| **Living without HIV + rifampin-resistant TB (Rif-R1**, n = 1,056**)** | | | | |
| **Sex+age** | 0.655 (0.600, 0.710) | ref | ref | ref |
| **Sex+age+** |  |  |  |  |
| Timika | 0.709 (0.656, 0.762) | 0.053 (0.007, 0.099) | 0.017 | 0.017 |
| PLI | 0.707 (0.656, 0.758) | 0.052 (0.011, 0.093) | 0.008 | 0.012 |
| ***Complete*** | 0.746 (0.695, 0.797) | 0.090 (0.035, 0.145) | 0.002 | 0.006 |

We estimated the prediction accuracy using Rif-S1 (n = 566) or Rif-R1 (n = 1,056) for models built with (A) individual radiological features, (B) PLI + cavitary information and (C) non-radiological + radiological features. We used a Monte Carlo cross-validation approach with 1,000 iterations of resampling (75:25), trained on the 75% and tested of the 25% of each iteration. At every iteration, we computed the difference between model AUCs (ΔAUC), and the number of observed differences that were ≤ 0 were divided by the total number of observations to assess statistical significance using a one-tailed empirical p-value approach [p-value = (#ΔAUC) ≤0/1,000]. We corrected for multiple hypothesis testing by controlling the Benjamini-Hochberg false discovery rate to <0.05. 𝝙AUC, Pval and FDR Pval were done compared to the sex+age model.

**Supplementary Table 5. Low- and high-risk group baseline characteristics for the test data when separated based on the optimal threshold for each logistic regression model.**

| **Characteristic** | **Sex+age+SG** | | **Sex+age+SG+PLI** | |
| --- | --- | --- | --- | --- |
|  | **low-risk (%)** | **high-risk (%)** | **low-risk (%)** | **high-risk (%)** |
| **Total** | 73 | 742 | 93 | 722 |
| **Female** | 64 (88) | 146 (20) | 76 (82) | 134 (19) |
| **Age at onset (years)** | 28 (22, 35) | 46 (37, 57) | 29 (24, 36) | 46 (37, 57) |
| **BMI ≤ 18 kg/m^2^** | 24 (33) | 280 (38) | 30 (32) | 274 (38) |
| **Prior TB** | 18 (25) | 242 (33) | 22 (24) | 238 (33) |
| **Extrapulmonary** | 1 (1) | 11 (1) | 1 (1) | 11 (2) |
| **Smoker** | 28 (38) | 493 (66) | 36 (39) | 485 (67) |
| **Alcohol use** | 12 (16) | 282 (38) | 12 (13) | 282 (39) |
| **Anemia** | 7 (10) | 134 (18) | 9 (10) | 132 (18) |
| **Other comorbidity** | 22 (30) | 375 (51) | 26 (28) | 371 (51) |
| **Smear grade (≥ 2+)** | 0 (0) | 259 (35) | 5 (5) | 254 (35) |
| **Rifampicin resistance** | 51 (70) | 479 (65) | 66 (71) | 464 (64) |
| **Effective TB regimen** | 62 (85) | 627 (85) | 77 (83) | 612 (85) |
| **Timika** | 8 (6, 27) | 45 (12, 72) | 8 (5, 20) | 48 (14, 72) |
| **PLI** | 8 (5, 14) | 20 (10, 40) | 7 (5, 10) | 22 (11, 41) |
| **Cavities** |  |  |  |  |
| Presence | 13 (18) | 339 (46) | 19 (20) | 333 (46) |
| Small | 13 (18) | 288 (39) | 19 (20) | 282 (39) |
| Medium | 2 (3) | 71 (10) | 0 (0) | 73 (10) |
| Large | 2 (3) | 40 (5) | 0 (0) | 42 (6) |
| Multiple | 3 (4) | 117 (16) | 1 (1) | 119 (16) |
| **Lymphadenopathy** | 15 (21) | 260 (35) | 18 (19) | 257 (36) |
| **Pleural effusion** | 4 (5) | 70 (9) | 5 (5) | 69 (10) |
| **Unfavorable outcome** | 3 (4) | 208 (28) | 3 (3) | 208 (29) |

**Supplementary Table 6. Machine learning model performance for prediction of PLI and Timika.**

| **Pre-training dataset** | **PLI (> 25%)** | | **Timika Score (56/140)** | |
| --- | --- | --- | --- | --- |
|  | **AUC (95% CI)** | **MAE (95% CI)** | **AUC (95% CI)** | **MAE (95% CI)** |
| **RSNA** | 0.77 (0.72-0.81) | 14.9 (13.7-16.1) | 0.71 (0.66-0.76) | 18.6 (17.2-20.2) |
| **NIH** | 0.78 (0.73-0.82) | 14.4 (13.1-15.6) | 0.74 (0.69-0.79) | 18.6 (17.2-20.0) |
| **PadChest** | 0.82 (0.78-0.85) | 12.7 (11.6-13.9) | 0.75 (0.71-0.80) | 16.6 (15.4-17.9) |
| **CheXpert** | 0.75 (0.72-0.79) | 14.3 (13.0-15.6) | 0.70 (0.63-0.76) | 17.8 (16.4-19.1) |
| **MMIC-CXR** | 0.75 (0.71-0.80) | 14.4 (13.1-15.6) | 0.74 (0.69-0.78) | 18.1 (16.8-19.5) |
| **MIMIC-CXR (CheXpert)** | 0.77 (0.74-0.80) | 14.3 (13.1-15.7) | 0.71 (0.66-0.76) | 18.7 (17.4-20.1) |
| **All** | 0.85 (0.82-0.88) | 11.7 (10.6-12.8) | 0.78 (0.73-0.83) | 15.8 (14.6-17.0) |

The classifiers’ performance was assessed using the Area Under the Receiver Operating Characteristic Curve (AUC-ROC), accuracy, precision-recall curve, F1 score, sensitivity, and specificity. Bootstrapping was used to estimate the AUC by resampling predictions and labels from the test set 100 times, generating a distribution of AUC scores with 95% confidence intervals. For performance evaluation in the Regression analysis, Mean Squared Error (MSE), Mean Absolute Error (MAE), and R-squared (R²) score were used. Bootstrapping was applied 500 times to generate a distribution of these metrics, and 95% confidence intervals were calculated.

Unfreeze last layer: trainable parameters 1,025.
